# A Systematic review and Network Meta-analysis of pharmaceutical interventions used to manage chronic pain

**DOI:** 10.1101/2023.02.23.23286360

**Authors:** Ash Shetty, Gayathri Delanerolle, Heitor Cavalini, Chunli Deng, Xiaojie Yang, Amy Boyd, Tacson Fernandez, Peter Phiri, Arun Bhaskar, Jian Qing Shi

**Affiliations:** University College London Hospitals NHS Foundation Trust; University College London, UK; Nuffield Department of Primary Care Health Sciences, University of Oxford, OX3 7JX, Oxford, UK; Southern University of Science and Technology, Shenzhen, 518055, China; Psychology Department, Faculty of Environmental and Life Sciences, University of Southampton, SO17 1BJ, Southampton, UK; Southern Health NHS Foundation Trust, SO40 2RZ, UK; Imperial College Healthcare NHS Trust, London, UK; School of Statistics and Mathematics, Yunnan University of Finance and Economics, China; National Centre for Applied Mathematics Shenzhen, China; University of Oxford, UK

## Abstract

**Background:** It is estimated 1.5 billion of the global population suffer from chronic pain with prevalence increasing with demographics including age. It is suggested long-term exposure to chronic could cause further health challenges reducing people’s quality of life. Therefore, it is imperative to use effective treatment options.

**Purpose:** We explored the current pharmaceutical treatments available for chronic pain management to better understand drug efficacy and pain reduction.

**Methods:** A systematic methodology was developed and published in PROSPERO (CRD42021235384). Keywords of opioids, *acute pain, pain management, chronic pain, opiods, NSAIDs, and analgesics* were used across PubMed, Science direct, ProQuest, Web of science, Ovid Psych INFO, PROSPERO, EBSCOhost, MEDLINE, ClinicalTrials.gov and EMBASE. All randomised controlled clinical trials (RCTs), epidemiology and mixed-methods studies published in English between the 1^st^ of January 1990 and 30^th^ of April 2022 were included.

**Data synthesis:** A total of 119 studies were included. The data was synthesised using a tri-partied statistical methodology of a meta-analysis (24), pairwise meta-analysis (24) and network meta-analysis (34).

**Limitations:** Small sample sizes, lack of uniformity with pain assessments and sub-optimal clinical trial designs were observed within the pooled data.

**Conclusion:** Chronic pain is a public health problem that requires far more effective pharmaceutical interventions with minimal better side-effect profiles which will aid to develop better clinical guidelines. The importance of understanding ubiquity of pain by clinicians, policy makers, researchers and academic scholars is vital to prevent social determinant which aggrevates issues.

## Introduction

Chronic non-cancer pain conditions are prevalent, highly debilitating and have high cost implications to health and social care. These conditions affect patients, their families and society at large, impacting approximately 20% of the global population^[1]^. The prevalence of pain conditions among females of all ages appears to be increasing^[2]^. Complexities around diagnosis and treatment of chronic pain conditions have meant that there is a paucity of standardised clinical guidelines that could potentially improve the clinical practice landscape, globally.

Convalescent periods for many chronically ill patients can be protracted and daunting. This may be especially true where pain medication has been used in the long term ^[3]^. Long-term exposures to chronic pain coincide with mental health and wellbeing issues, exacerbating patient-reported outcomes such as sleep disturbances, depression, dependence and morbidities such as myalgia, lethargy and fatigue^[4]^. Better understanding of long-term implications requires consideration of “life-course approaches” and at present, this could evolve further within pain medicine epidemiology ^[5]^.

The increase in chronic pain conditions contributes to higher healthcare costs towards clinical management of patients, and also reduced levels of productivity for employers ^[6]^. This may be partly due to increases in opioid use within this population of patients, often reducing their capacity to conduct normal working hours. Current clinical guidelines recommend non-invasive pain management options as a first-line treatment among non-cancer patients in particular, although overdose, dependency and mortality due to opioid use has consistently increased over time ^[7]^. It was reported that global opioid use has doubled between 2011 and 2003 to 2011 and 2013 to 7.35 billion daily doses per year^[6]^. These figures were attributed to North America and Europe largely and could reflect the increase in the population ^[7]^. Attrition data with regards to differentiating those with recreational use and those on medical prescription for a long-term conditions during this period remains scarce.

The World Health Organisation’s (WHO) analgesic framework could be a useful model to consider while determining the rates and prevalence of pain management methods including opioid use among those with and without cancer pain^[8]^. It is particularly important to develop evidence-based guidelines specific to each condition, with flexible pain medication use as a single regimen or a combination of treatments that could improve the overall quality of life of these patients^[9]^. The premise to increase the strength and frequency of pain medications is in general based on disease burden i.e. progression of symptoms and patients reported symptoms. Pain inferences in particular could play a vital role in the use of analgesics ^[10]^. The primitive consciousness and the link to mental health is yet another factor that would exacerbate pain medication use. Additional approaches, for example using a cognitive behavioural-based therapeutic protocol with pharmaceutical management can benefit patients ^[10]^. However, the benefits of these approaches remain unclear due to a paucity of evidence from both clinical trials and real-world study evidence. We have designed the POP project as the initial step to conduct exploratory work on pharmaceutical management of chronic pain. With the rising need for comparative effectiveness research, increasingly more systematic reviews focus on evaluating the relative efficacy and acceptability of drugs and therapeutic interventions^[11]^. However, some of the interventions for long-term conditions are not available for clinical practice and there are several options with varying efficacy even within a specific class of interventions^[12]^.

## Methods

We developed a wide systematic methodology and published this as a protocol with multiple research questions in the first instance in PROSPERO (CRD42021235384). Data from studies meeting the inclusion criteria were extracted and Pairwise Meta-Analysis with random and fixed effects models was carried out. Pooled mean difference (MD) together with 95% confidence intervals (CIs) are reported overall and for sub-groups. By combining the direct and indirect comparisons between different interventions, Network Meta-Analysis was conducted to explore the relative treatment effects among all the drugs included in our analysis.

### Aims

The aims of the study was to explore the prevalence of treatments of effects in chronic pain based on pharmaceutical treatments.

### Search strategy

The search strategy used key words of *chronic pain, opioids, acute pain, pain management, opiods, NSAIDs, analgesics* across multiple databases (PubMed, Science direct, ProQuest, Web of science, Ovid Psych INFO, PROSPERO, EBSCOhost, MEDLINE, ClinicalTrials.gov and EMBASE).

### Eligibility criteria

All randomised controlled clinical trials (RCTs), epidemiology and mixed-methods studies reporting the use of pain medication for non-cancer chronic pain conditions published in English between the 1^st^ January 1990 and 30^st^ April 2022 were included. Opinions, commentaries and editorials were excluded.

## {PRISMA diagram}

### Data extraction

Participants included in the study populations were those who were experiencing chronic pain. All studies reporting drug efficacy associated with chronic pain were extracted by way of the interventions, measures of tool and numeric results. An extraction template specific to the objectives of the study was developed to gather a wider dataset with vital data for statistical analysis. The number of studies was the number of independent RCTs included in analysis, however sub-studies were extracted from the same clinical trials with different duration periods. The results of different stages in one designed study can be regarded as new sub-studies as new rows in data analysis.

Data was extracted by two investigators and any disputes for eligibility was discussed and agreed with the Chief Investigator of the study. All studies included within the analyses were independently reviewed.

### Outcome measures

Outcomes were reported as mean, median, standard deviation and confidence intervals. Mean and standard deviation were extracted as the main outcomes including pre-treatment pain scores at baseline, post-treatment pain scores and pain score changes of each group. Multiple pain assessments for confirming a clinical diagnosis, severity and progression of chronic pain were identified. These include VAS (visual analogue scale, 0-10 or 0-100), NRS (11-point numeric rating scale, 0-10), BPI (Brief Pain Inventory interference scale, 0-10), MPQS (McGill Pain Questionnaire-Short Form (Sensory and Affective subscales, VAS intensity measure, 0-10), VRS (verbal rating scale, 0-10), NIH-CPSI (National Institutes of Health Chronic Prostatitis Symptom Index, pain scores, 0-21), PI (pain intensity on a 20-point scale, 0-20).

As most widely used tools for assessing pain such as VAS, NRS, VRS, use a 11-point numeric rating scale from 0 to 10, the following standardisation formula was used to unify all pain scores into the same scale:

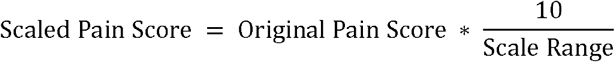

As all outcomes of interest were continuous, the calculation based on pain scores was performed by using mean differences (MD) with a 95% confidence interval (CI) to report the effects between the group comparisons.

### Exposures

The exposures of interest were selected based on the key features of pharmacological management used to treat non-cancer chronic pain, including and not limited to a pain condition being the primary or the secondary condition. Neurological and psychological symptoms leading up to the use of pharmaceutical use within the included population were also considered.

### Statistical analysis plan

A meta-analysis, pairwise meta-analysis (PMA) and Network meta-analysis (NMA) were used to compare all treatments used in managing chronic pain. The fundamental difference between them is that PMA produces only one estimate of pooling effects from the selected pair of interventions, while NMA produces multiple comparative estimates of pooling effects by connecting all alternative interventions. ^[14]^

We incorporated direct and indirect treatment comparisons within the NMA providing greater statistical precision compared to a PMA ^[20]^. Rankings of a set of drugs or combined interventions for assessing chronic pain with respect to their efficacy was calculated based on the netowkr models. Homogeneity and Consistency were tested to see if the assumptions in NMA were violated. NMA was conducted with 2-arm and multi-arm studies including more types of Pharmaceutical interventions. PMA was conducted with 2-arm studies with the placebo as the control group while a pharmaceutical intervention as the experimental group. The overall pharmaceutical efficacy of extracted studies was produced by pooling all treatment effects. PMA was also used on studies with the same drug as the treatment group to see the specific drug efficacy.

I^2^and p-value were commonly used to detect statistical heterogeneity. A value of I^2^ larger than 50% with a much smaller p-value indicates strong heterogeneity. Correspondingly, I^2^ less than 50% with a large p-value indicates fairly weak heterogeneity ^[16]^. A random effects model was chosen when there was high heterogeneity, whereas a fixed effects model was used if weak or no heterogeneity was detected ^[17]^. Due to the presence of high heterogeneity, subgroup analyses were carried out to identify the sources. To assess the robustness of the pooled results within the PMA, a sensitivity analysis was completed. Publication bias was evaluated with funnel plots and Egger tests. The statistical analyses were produced by R and packages were used to provide outputs in compliance with best practice and reporting guidelines ^[18]^.

## Results

Of the 119 systematically included studies (Table 1) with 17,708 participants, 24 were used in the meta-analysis. The 24 studies were placebo-controlled RCTs with a primary endpoint of efficacy. Of the 119 studies, 34 were used in the NMA to build a connected network.

**Table 1.**
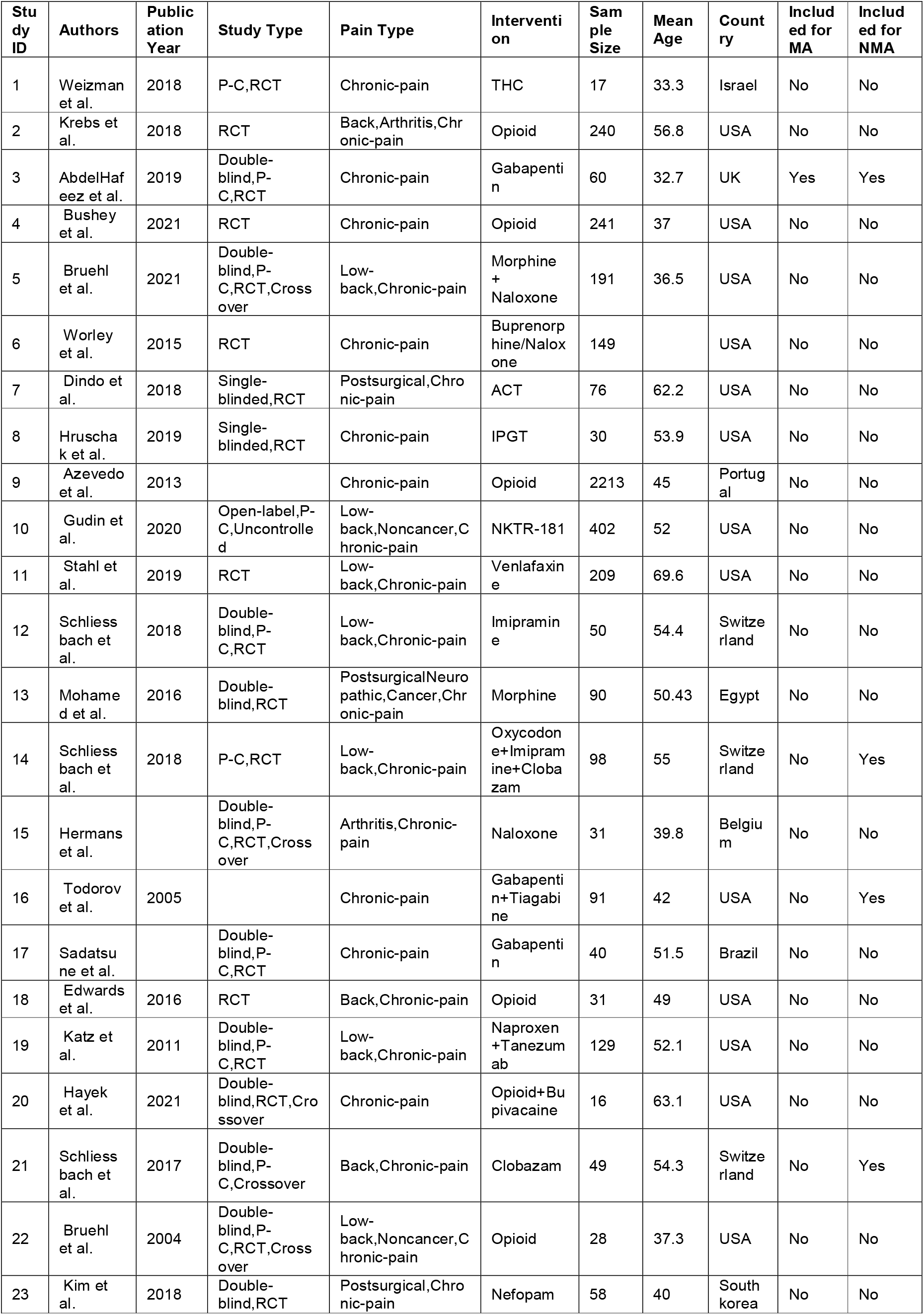
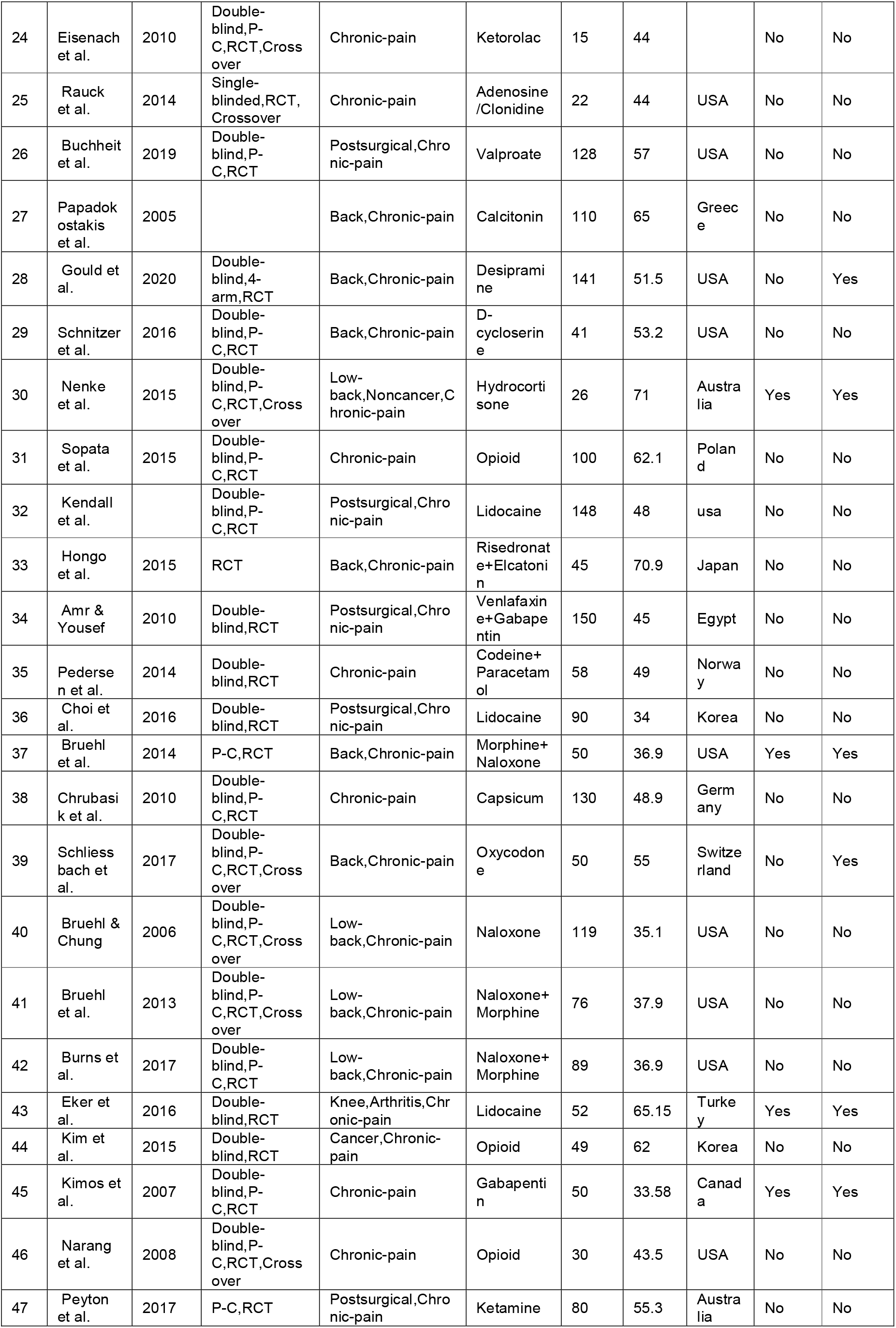
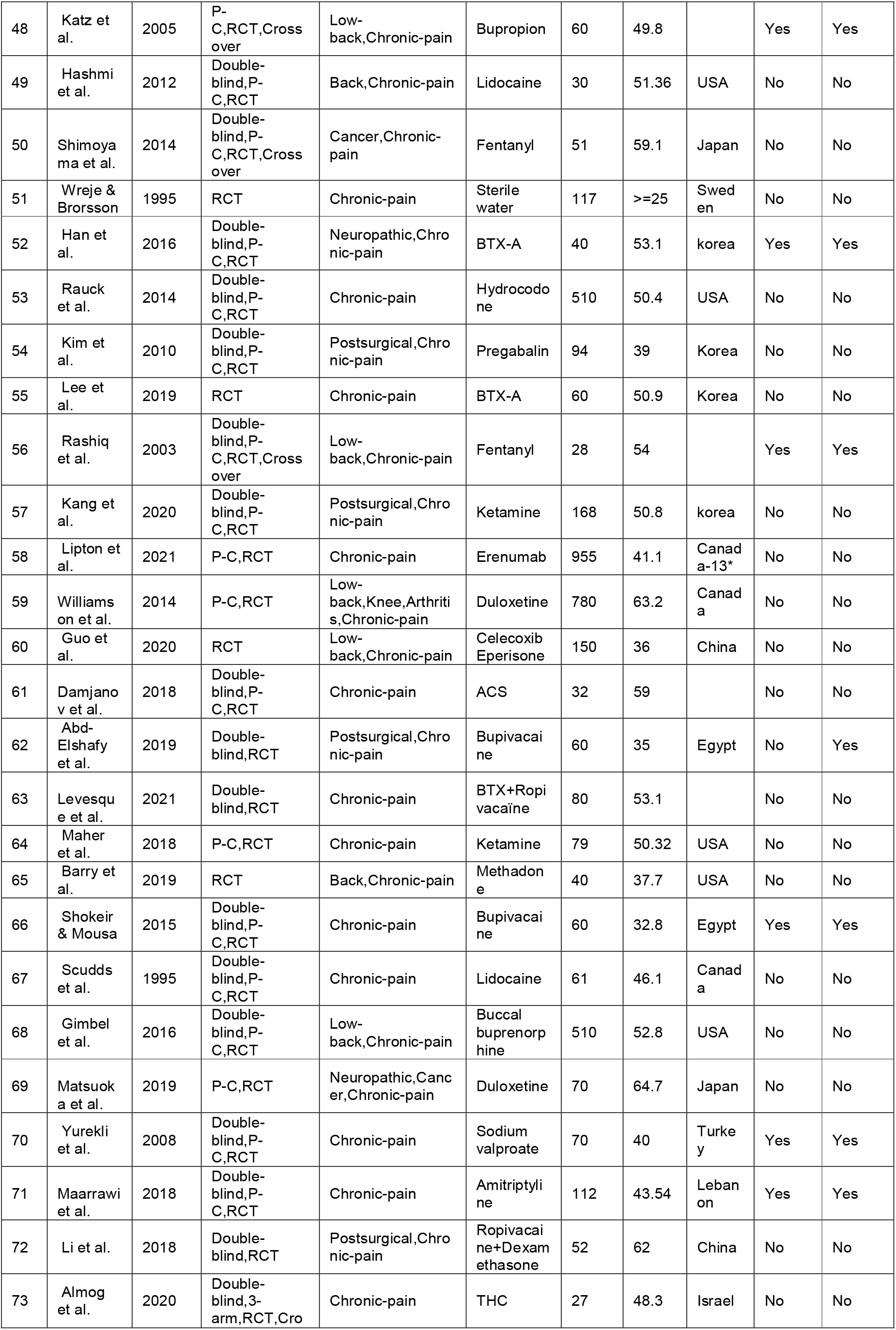
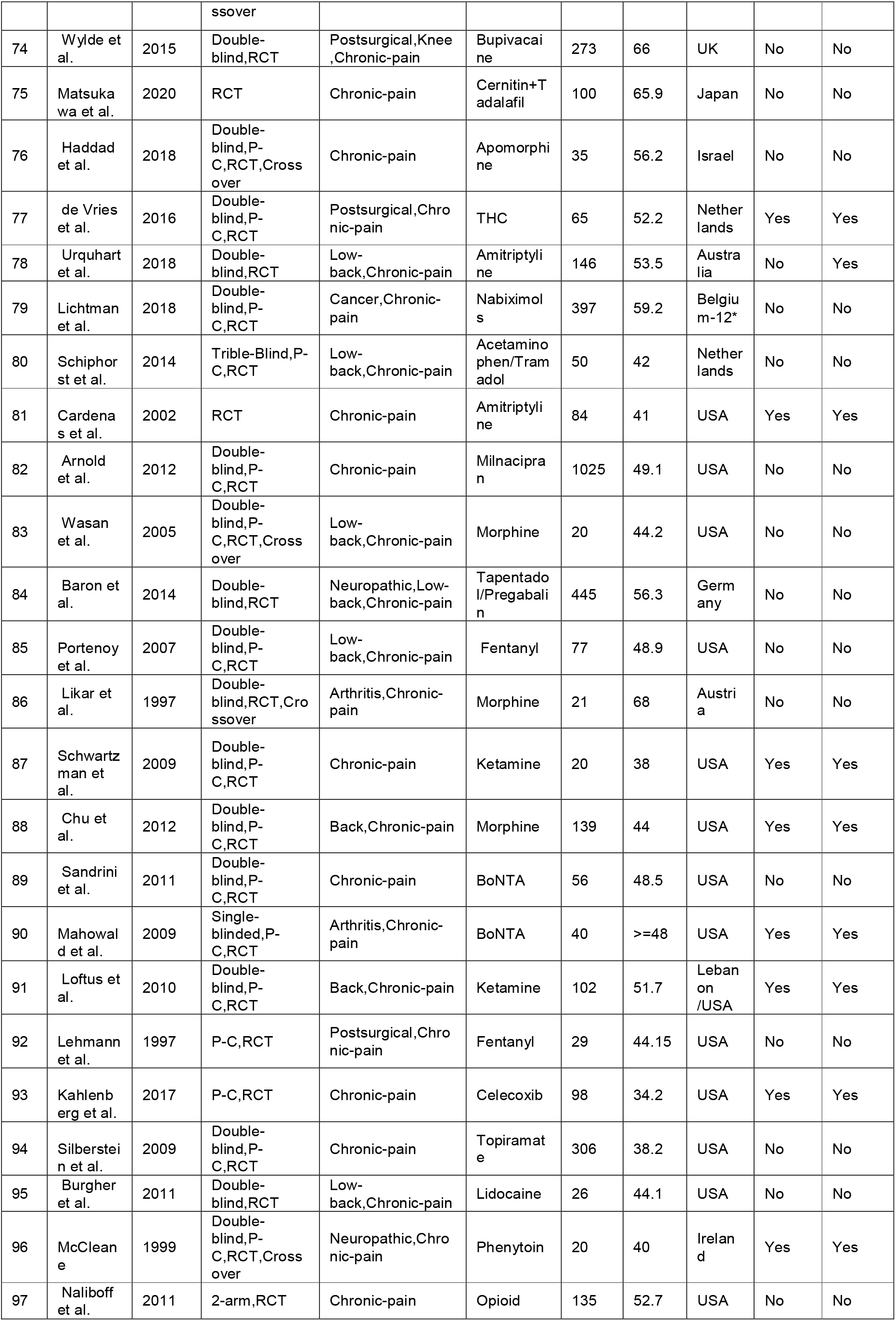
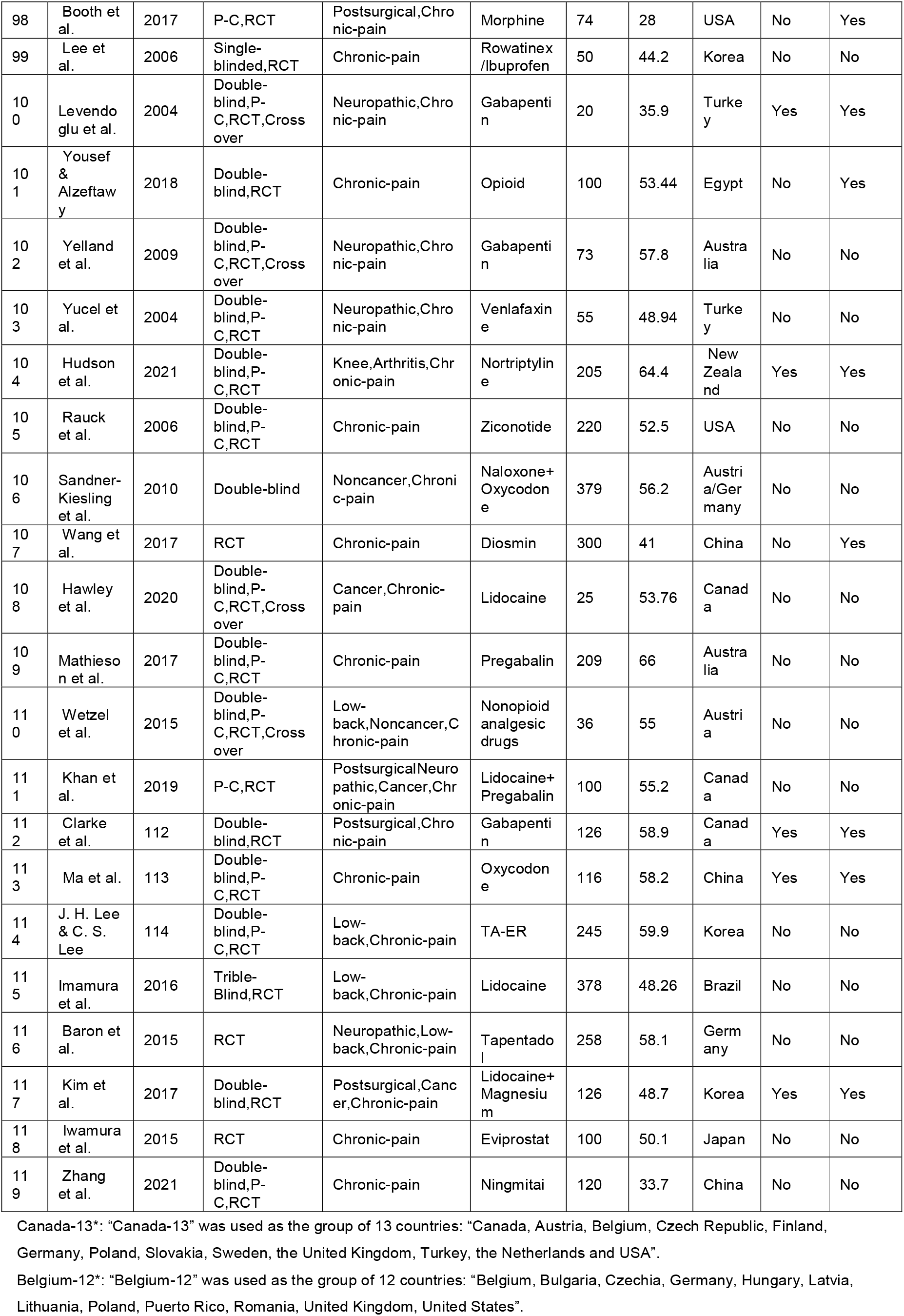
Characteristics of the studies included in systematic review

Opioids (Table 2) were most commonly tested in 32 (26.89%) studies enrolling 5518 (31.16%) participants, where *Morphine, Oxycodone* and *Fentanyl* were prominently tested. *Lidocaine, Naloxone* and *Gabapentin* were the most frequently tested non-opioid drugs for chronic pain. The most common pain among chronic pain patients were lower back pain, which was explored in 26 (21.85%) studies with a pooled sample of 4626 (26.12%) while 13 studies reported chronic back pain among 1068 (6.03%) participants. The following pain types are post-surgical pain and neuropathic pain with 19 (15.97%) and 10 (8.4%) studies involved to test the efficiency of NSAID drugs on patients.

**Table 2.** Summary of drug and pain types included in systematic review

| Summary of drug and pain types included in studies systematic review |  |  |  |
| --- | --- | --- | --- |
| Classes | Drug types | Studies (Number, %) | Participants (Number, %) |
| Opioids<br>32 (26.89%) | Oxycodone | 4 (3.36%) | 643 (3.63%) |
|  | Fentanyl | 4 (3.36%) | 185 (1.04%) |
|  | Methadone | 1 (0.84%) | 40 (0.23%) |
|  | Morphine | 9 (7.56%) | 750 (4.24%) |
|  | Buprenorphine | 2 (1.68%) | 659 (3.72%) |
|  | Codeine | 1 (0.84%) | 58 (0.33%) |
|  | Other Opioids | 11 (9.24%) | 3183 (17.97%) |
| Nonopioids | Naloxone | 8 (6.72%) | 1084 (6.12%) |
|  | Gabapentin | 8 (6.72%) | 610 (3.44%) |
|  | Lidocaine | 10 (8.4%) | 1036 (5.85%) |
|  | Ketamine | 5 (4.2%) | 449 (2.54%) |
|  | Amitriptyline | 3 (2.52%) | 342 (1.93%) |
|  | Bupivacaine | 4 (3.36%) | 409 (2.31%) |
|  | THC | 3 (2.52%) | 109 (0.62%) |
|  | Others | 54 (45.38%) | 9251 (52.24%) |
| Categories | Pain types | Studies (Number, %) | Participants (Number, %) |
| Chronic Pain | Postsurgical | 19 (15.97%) | 1987 (11.22%) |
|  | Neuropathic | 10 (8.4%) | 1171 (6.61%) |
|  | Low Back | 26 (21.85%) | 4626 (26.12%) |
|  | Cancer | 8 (6.72%) | 908 (5.13%) |
|  | Back (Except Low-Back) | 13 (10.92%) | 1068 (6.03%) |
|  | Knee | 4 (3.36%) | 1310 (7.4%) |
|  | Arthritis | 7 (5.88%) | 1369 (7.73%) |
|  | Pelvic | 4 (3.36%) | 300 (1.69%) |
|  | Other Chronic Pain | 42 (35.29%) | 8578 (48.44%) |

**Table 5.** Egger test results for studies used in PMA

|  |  |  |  |  |
| --- | --- | --- | --- | --- |
| Test result: | t = 1.24, df = 29, p-value = 0.2247 |  |  |  |
|  | bias | se.bias | intercept | se.intercept |
| Sample estimates: | 1.49 | 1.2 | -1.593 | 0.3737 |

Meta-analysis of mean difference of pain scores were applied to 24 studies with a sample of 2546 participants, producing a pooled mean difference (MD) of -0.89 (95% CI = [-1.31, -0.47]). There was a significant difference between chronic pain scores of patients taking NSAIDs compared to a placebo. Averagely, 0.89 point (0-10 scale) of pain reduction was observed based on the random effects model. A significant statistical drug efficiency was observed with BTX-A and Ketamine. A negative pooled mean difference was determined between BTX-A and Ketamin versus a placebo with a pain reduction of 0.98 -1.26 based on a –10 scale, respectively. Similar statistical results were not obserbed with other drugs in comparison to a placebo.

With the common comparator as “*placebo*”, the connected network included 34 studies, 52 pairwise comparisons, 32 interventions and 29 study designs. Gabapentin had a significant mean difference equaling to -1.49 (95% CI = [-2.76, -0.23], p-value < 0.05). Most interventions had a negative mean difference compared to a *placebo*, but a 95% CI covering 0 indicated insignificant effects for reducing pain. The results within the network were more conservative with the combination of direct and indirect evidence indicating most pharmaceutical interventions selected might have benefited from the “*placebo effect*”.

### Pairwise Meta-Analysis (PMA)

The PMA included 24 studies with pairwise comparisons between drugs and a placebo. The experimental and control group comprised of *“Amitriptyline”, “BTX-A”,”Gabapentin”, “Ketamine”, “Lidocaine”, “Morphine”, “Naloxone”* and a *placebo*, respectively. A single study reported “*Fentanyl”, “Ningmitai”, “THC”, and “Oxycodone*”.

### PMA for Baseline Pain Score

The PMA was used to test baseline pain score differences between the experimental and control group in 18 studies which comprised of a total sample of 1,691 participants. The experimental and control groups comprised of 837 and 854 participants, respectively, with a pooled mean difference (MD) of -0.02 (95% CI = [-0.13, 0.08]). The 95% CI was 0 and therefore, no statistically significant difference between baseline pain scores of two groups. [Figure 1]. A weak statistical heterogeneity of 15% of *I*^2^ (p = 0.26) was determined. This combined with the statistical insignificance indicates the randomisation of was completed accurately and that it is scientifically justifiable to use the post-treatment pain scores directly as the outcomes to evaluate treatment effects.

**Figure 1.**
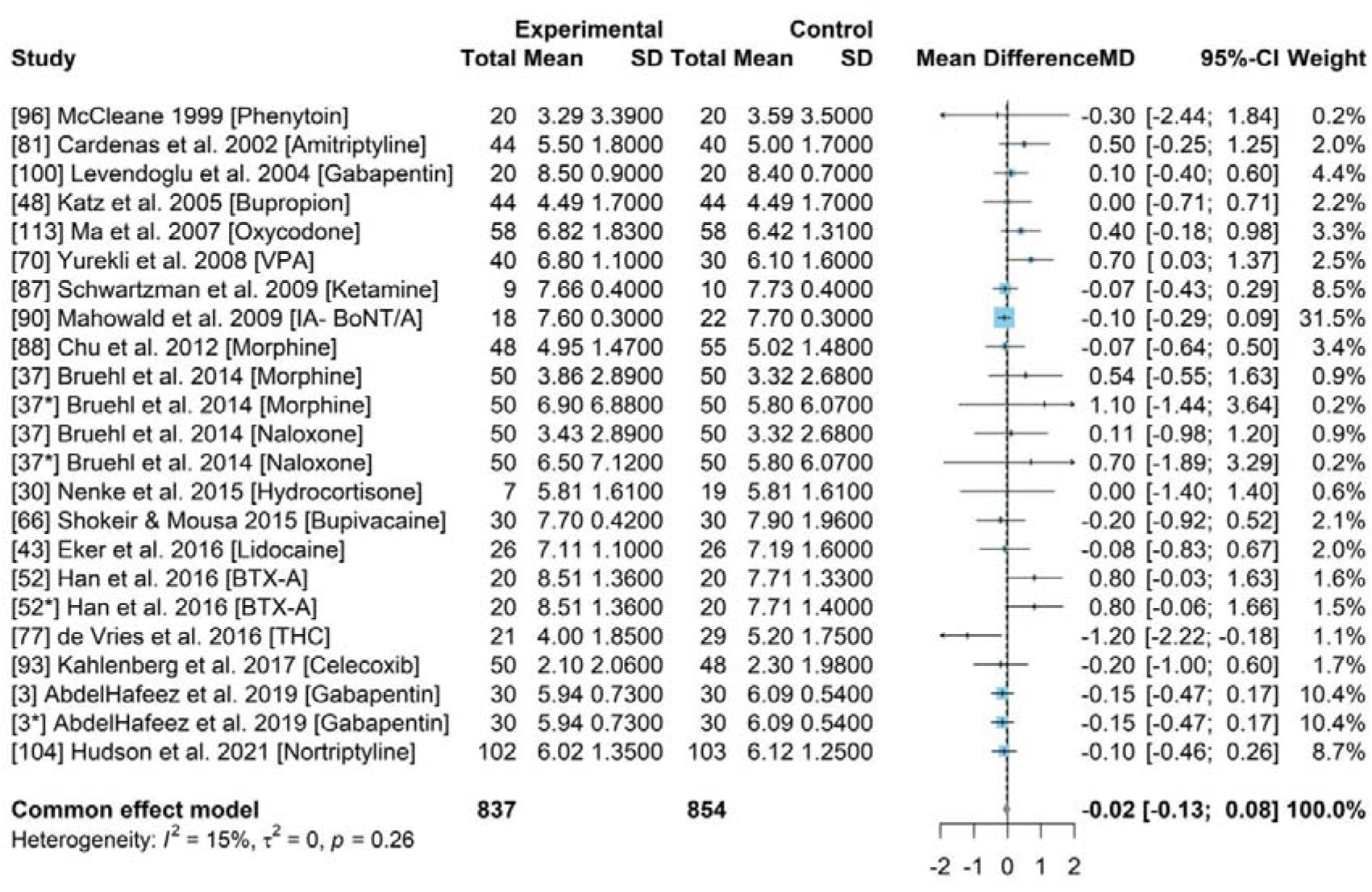
Forest plot for the baseline pain scores of experimental group and control group across 18 studies.

### PMA for drug efficacy between NSAID compared to a placebo

This PMA included 24 studies [Figure 2] with 2418 participants, with a MD of -0.89 (95% CI = [-1.31, -0.47]). The experimental and control group comprised of 1219 amd 1199, respectively. A significant statistical heterogeneity of 92% of *I*^2^ (p-value <0.01) was identified. Mean difference (MD) was calculated to assess if there is statistically significant difference of post-treatment pain scores between experimental group and control group. The 95% CI was less than 0 which indicated a significant treatment effect with a reduction in pain by 0.89-point (0-10 scale) compared to those who were given a placebo.

**Figure 2.**
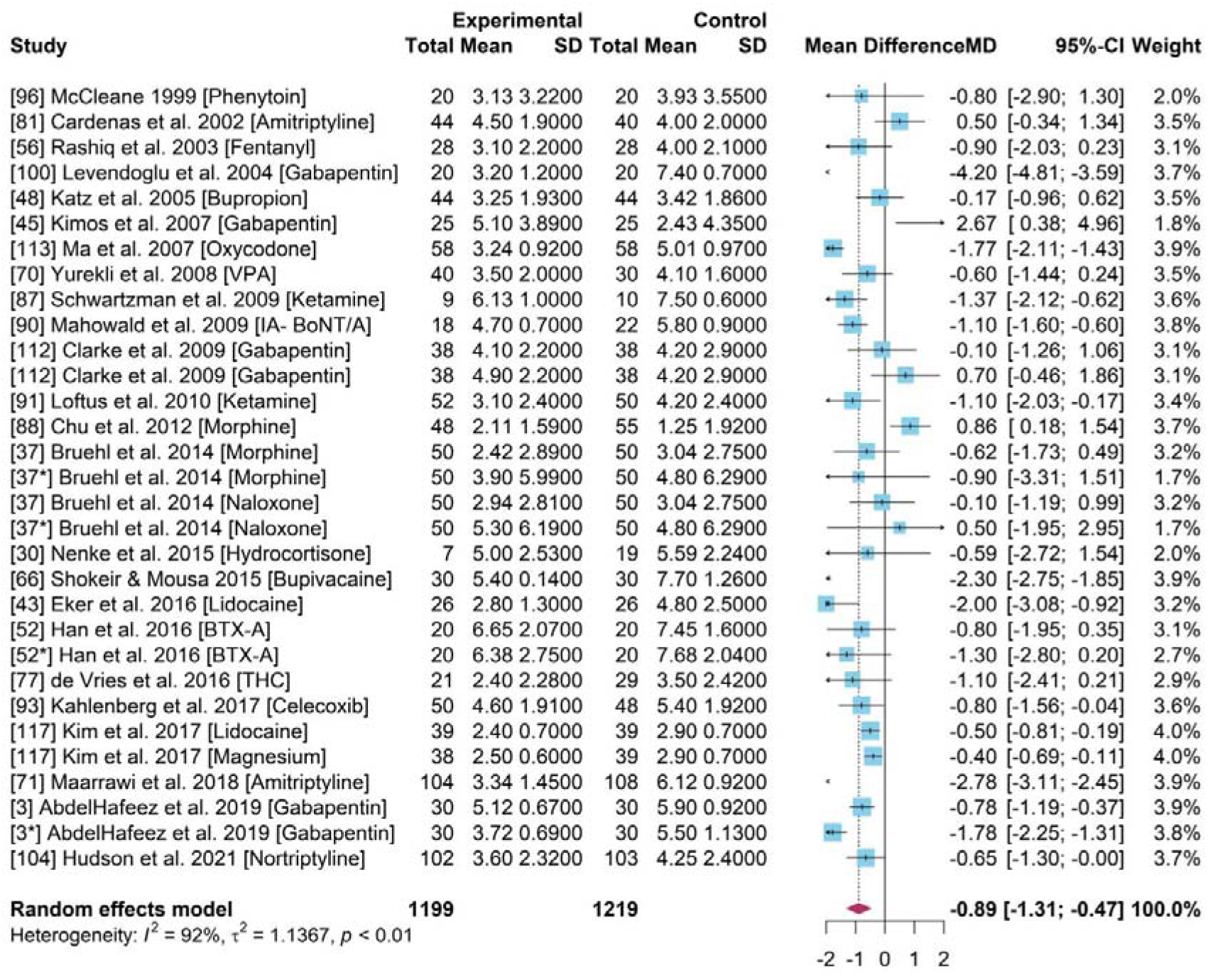
Forest plot for the pain scores of experimental group and control group across 24 studies testing all NSAID drugs (including some unnamed Opioids drugs).

### Meta-Analyses

A statistically low heterogeneity of 0% of *I*^2^ (p-value > 0.5) was identified among studies with *BTX-A, Ketamine* and *Naloxone* [Figure 3b and 3d]. *BTX-A* [Figure 3b] and *Ketamine [*Figure 3d] indicated statistically significant drug efficacy of -1.07 [-1.51, -0.64] and -1.26 [-1.85, -0.68], respectively. The treatment efficiency compared to the placebo had a 1 point pain reduction within a 0-10 evaluation scale. Ketamine demonstrated optimal efficacy with a 1.26 point pain reduction on average.

**Figure 3a.**
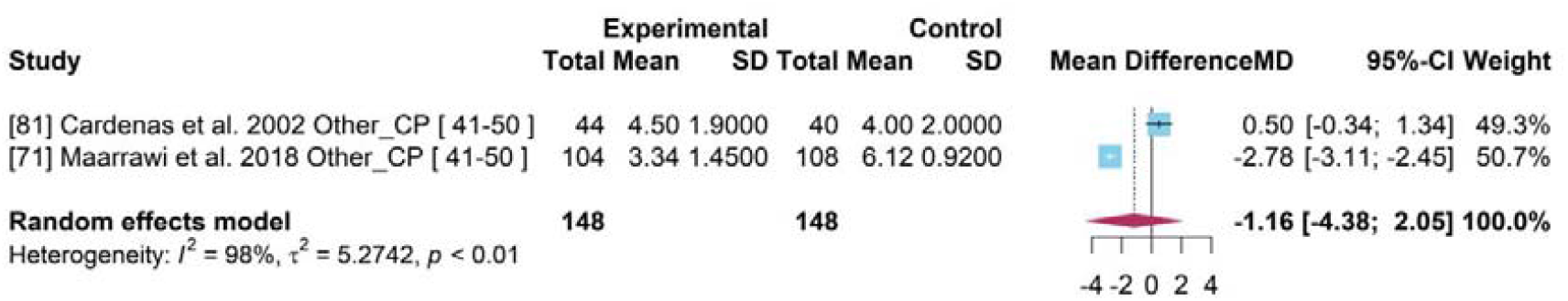
Forest plot for drug efficiency of Amitriptline.

**Figure 3b.**
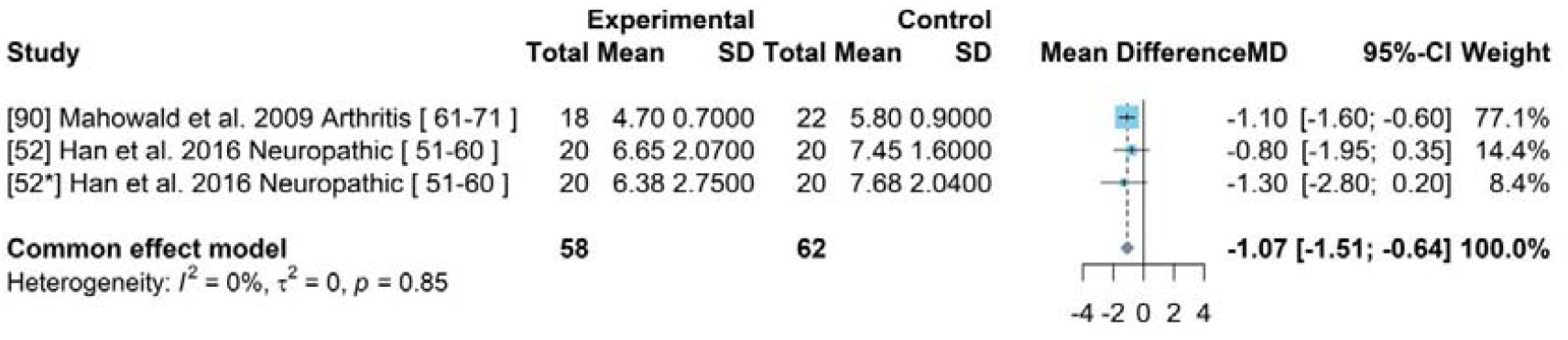
Forest plot for drug efficiency of BTX-A.

**Figure 3c.**
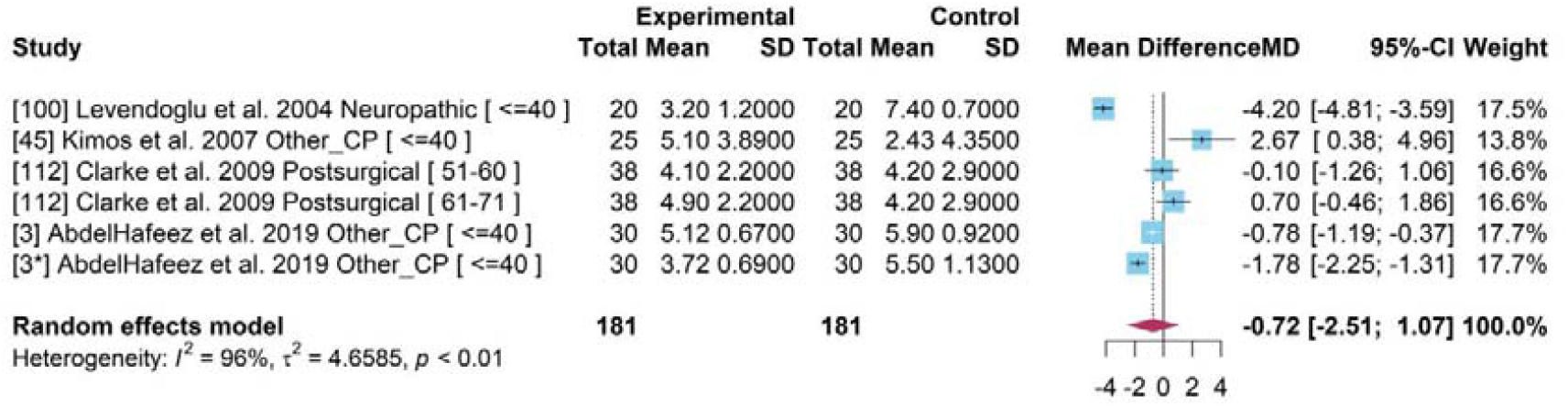
Forest plot for drug efficiency of Gabapenin.

**Figure 3d.**
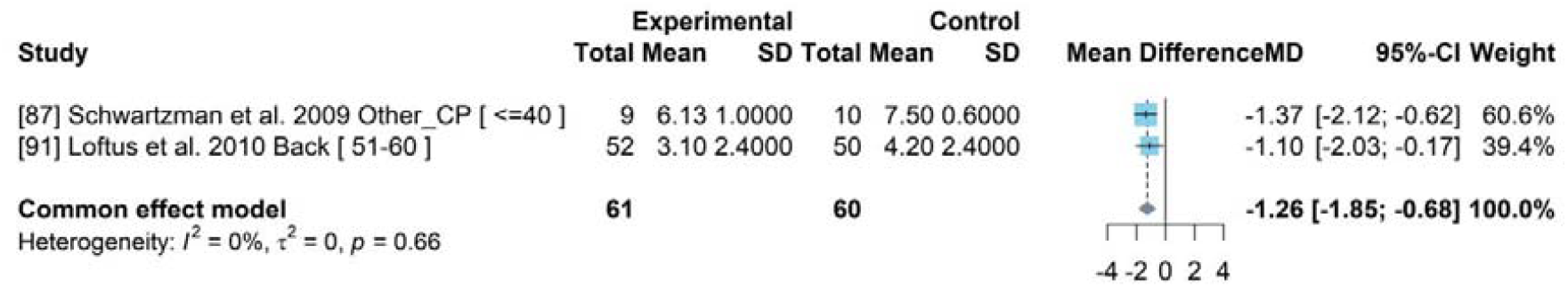
Forest plot for drug efficiency of Ketamine.

**Figure 3e.**
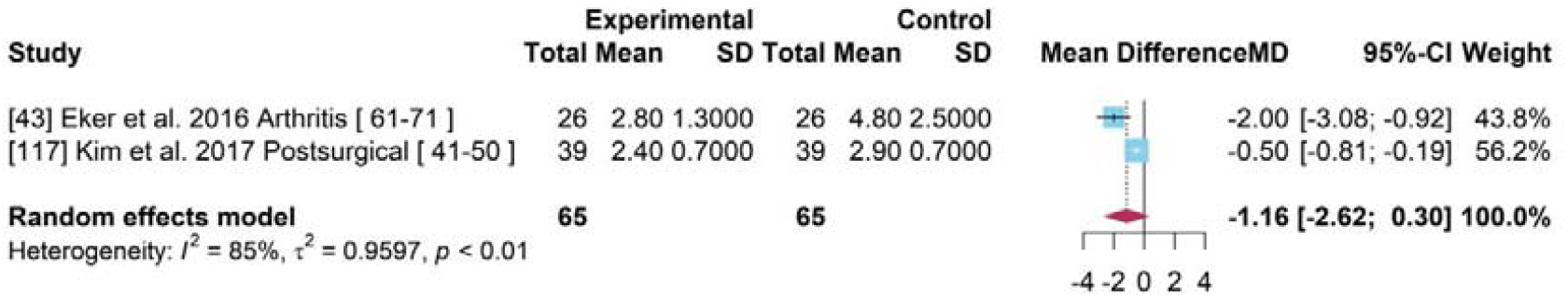
Forest plot for drug efficiency of Lidocaine.

**Figure 3f.**
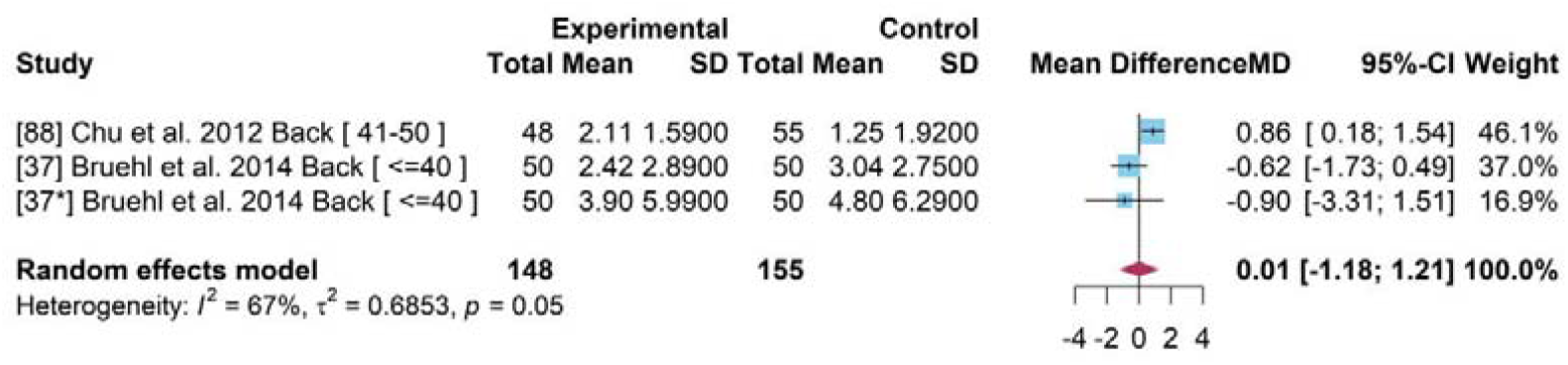
Forest plot for drug efficiency of Morphine.

**Figure 3g.**
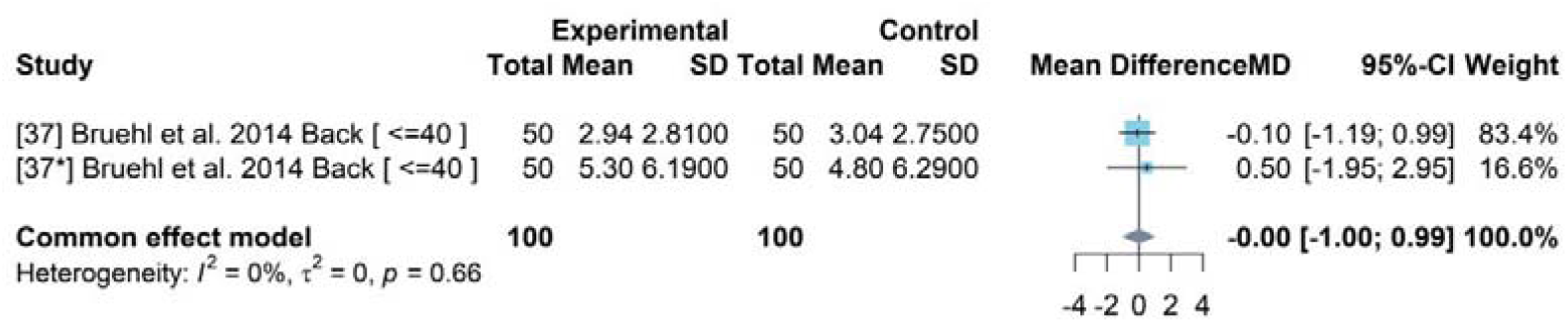
Forest plot for drug efficiency of Naloxone.

The PMA for *BTX-A* (Figure 3b) and *Naloxone* (Figure 3g) showed a low heterogeneity as the data was pooled from a single study.

Studies on *Amitriptyline, Gabapentin, Lidocaine and Morphine* had a high heterogeneity and a statistically insignificant drug efficacy (Figure 3a, c, e, f). The mean difference of 95% CI was 0 indicating an insignificant treatment difference between the drugs and placebo based on the random effects model.

### Opioids drugs

A meta-analysis was conducted with 4 studies with Opioids [Figure 4]. A pooled MD of -0.65 and a 95% CI [-1.67, 0.37] was determined indicating an insignificant treatment effect of opioids drugs compared to a placebo. A statistically sginficant heterogeneity of 92% of *I*^2^ (p-value < 0.01) was identified.

**Figure 4.**
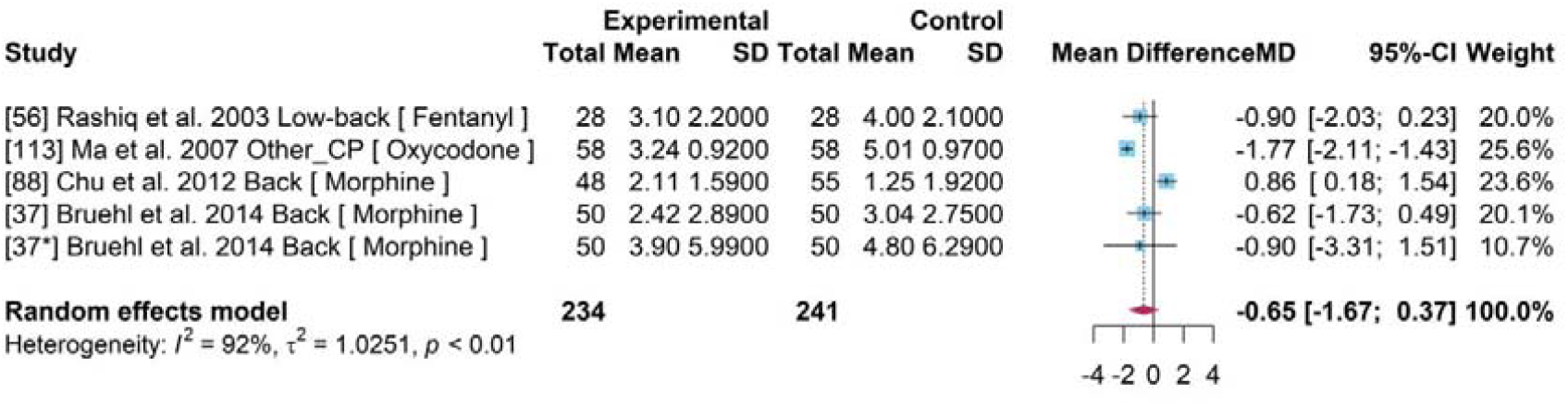
Forest plot for drug efficiency of Opioids drugs*.

### Network Meta-analysis (NMA)

A NMA [Figure 5] was completed for 34 studies. The nodes correspond to each intervention included within the network where the interventions with direct comparisons are linked with a line. The thickness of lines corresponds to the number of trials evaluating the comparison. A connected network was built based on the *placebo* which was mostly *Tolterodine* based on the original studies. The evaluations between interventions were supported by direct comparison and indirect comparison

**Figure 5.**
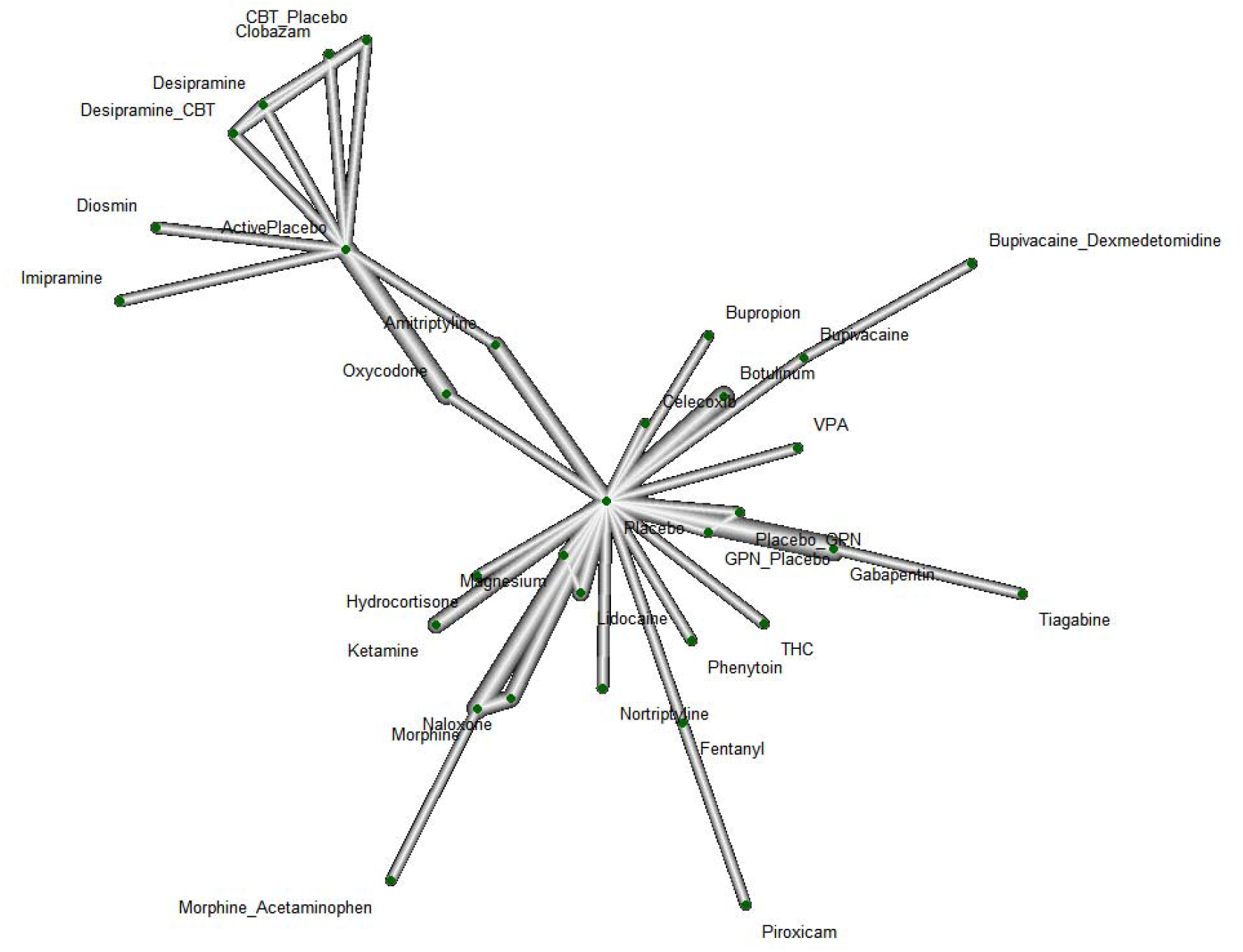
Network plot where Placebo was the reference group with 34 studies and 32 interventions. Opioids drugs*: including Fentanyl, Oxycodone, Morphine and other unnamed Opioids drugs.

In the network with the placebo as the reference group, *Gabapentin* [Figure 6] comprised of a MD equaling to -1.49 (95% CI = [-2.76, -0.23], p-value < 0.05) indicating a significant effect on reducing chronic pain and direct comparisons were made using 4 studies [Figure 7a]. The pooled MD of *Botulinum* and *Ketamine* were -1.06 and -1.24, respectively. These were similar to the results in the PWA, but their 95% CI was 0 therefore showed insignificant effect on pain reduction compared to a placebo. Most combined interventions had a negative MD compared to a placebo with a 95% CI of 0 indicated statistically insignificant results for reducing pain.

**Figure 6.**
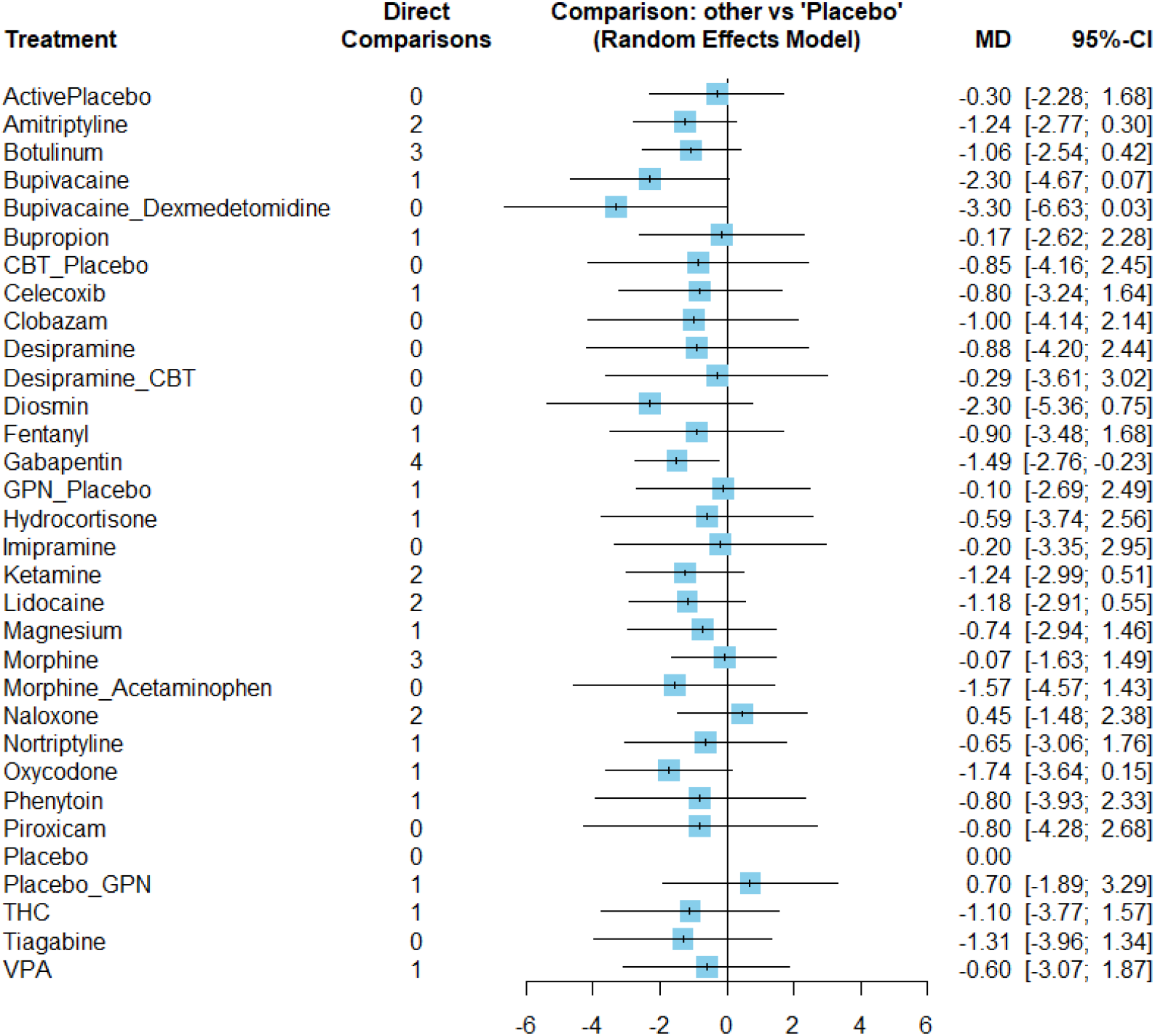
Forest plot for intervention efficiency compared to Placebo in NMA

**Figure 7a.**
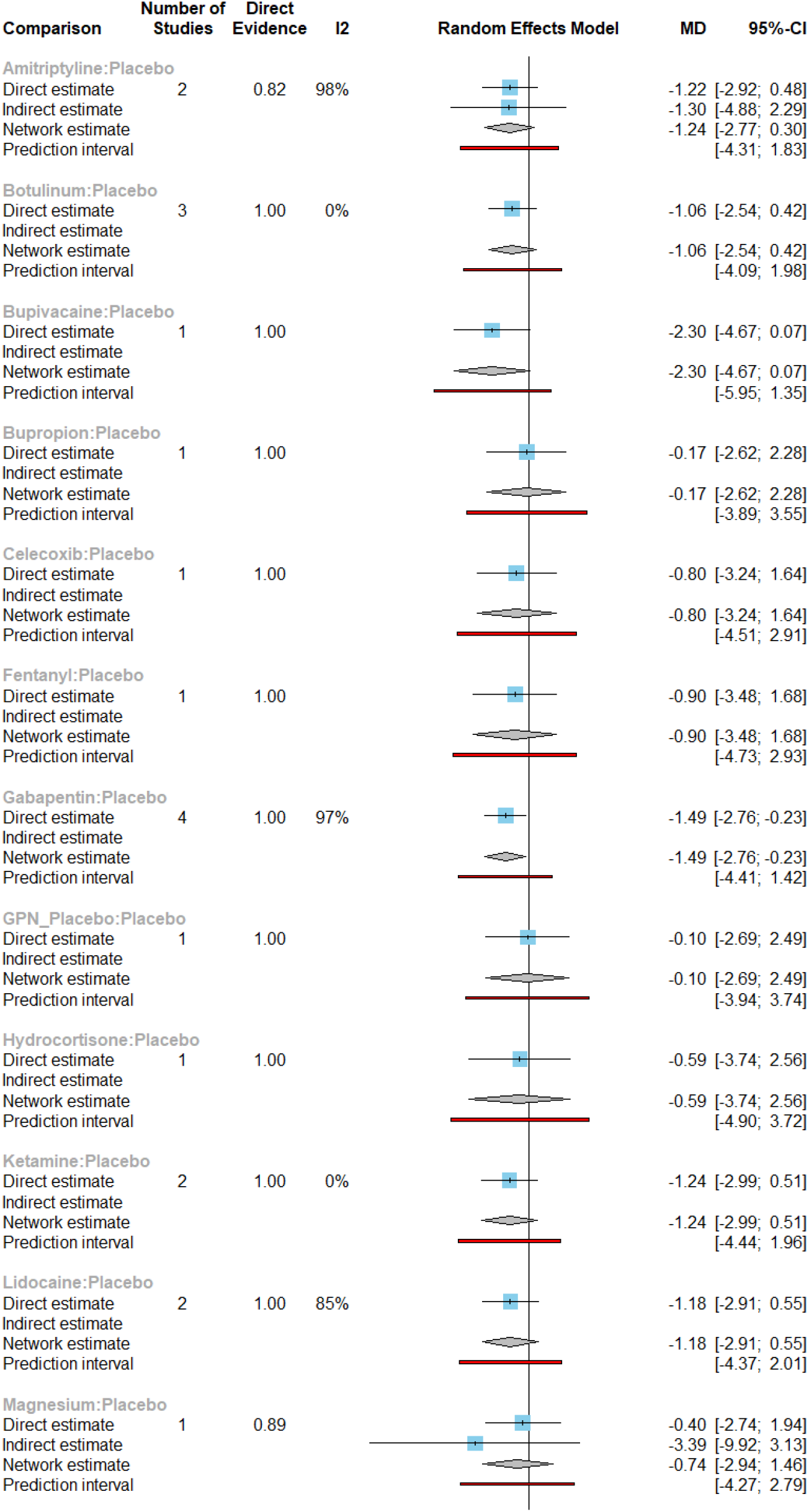
Forest plot for intervention efficiency compared to Placebo in NMA with detailed direct and indirect comparisons

**Figure 7b.**
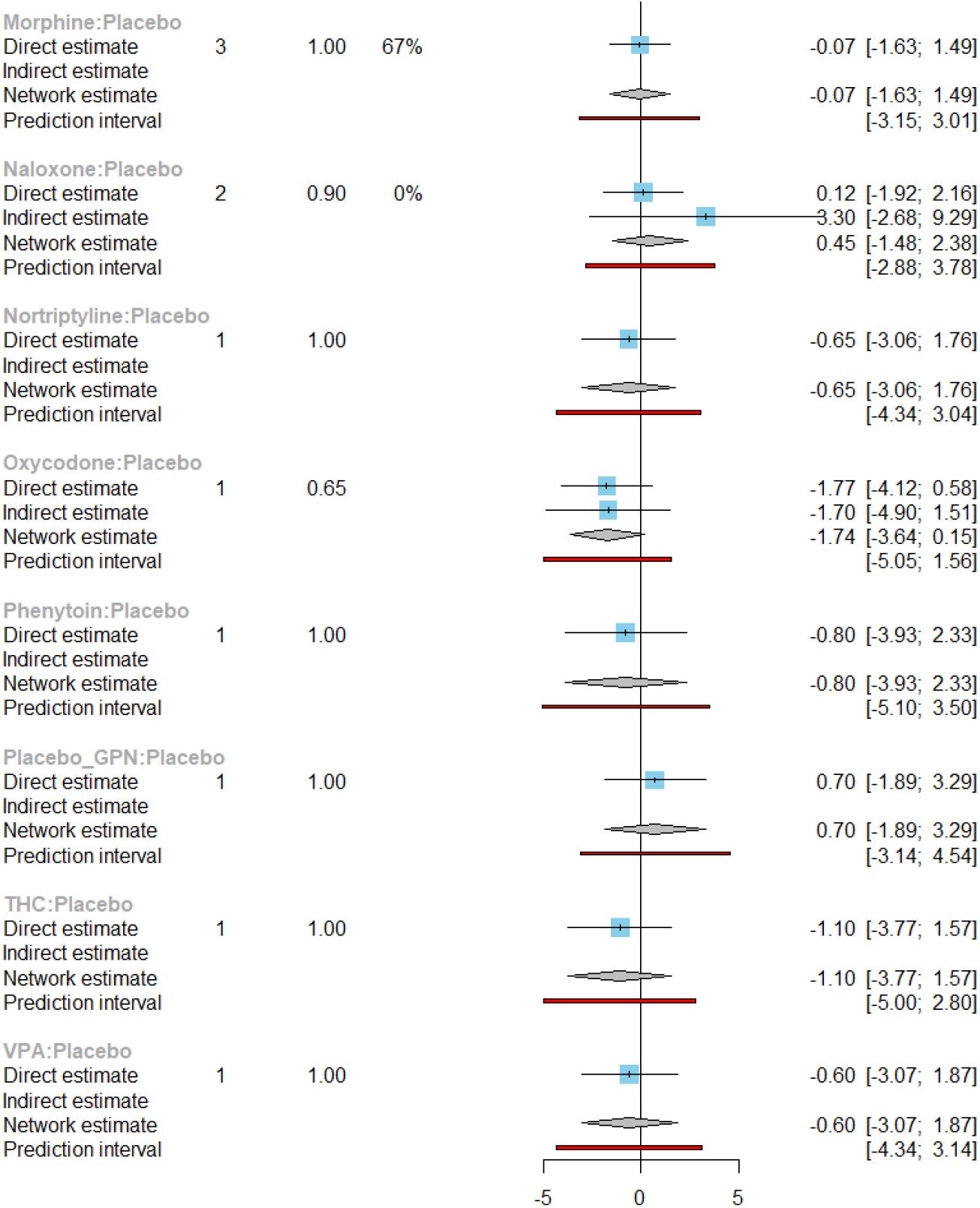
Forest plot for intervention efficiency compared to Placebo in NMA with detailed direct and indirect comparisons

*Imipramine, Diosimin, Desipramine, Clobazam, Piroxicam* and *Tiagabine* had not been directly compared to a placebo based on the identified data therefore the comparative treatment effected between them and a placebo was not possible to complete.

### Subgroup Analysis

A subgroup analyses was conducted for 24 studies within the meta-analysis to explore the sources of heterogeneity and unbiased estimation based on age, pain type, period and geographical location [Figure 8]. The sub-group analysis for pain type, time period and geographical location can be found in the supplementary file whilst average age is shown below.

**Figure 8.**
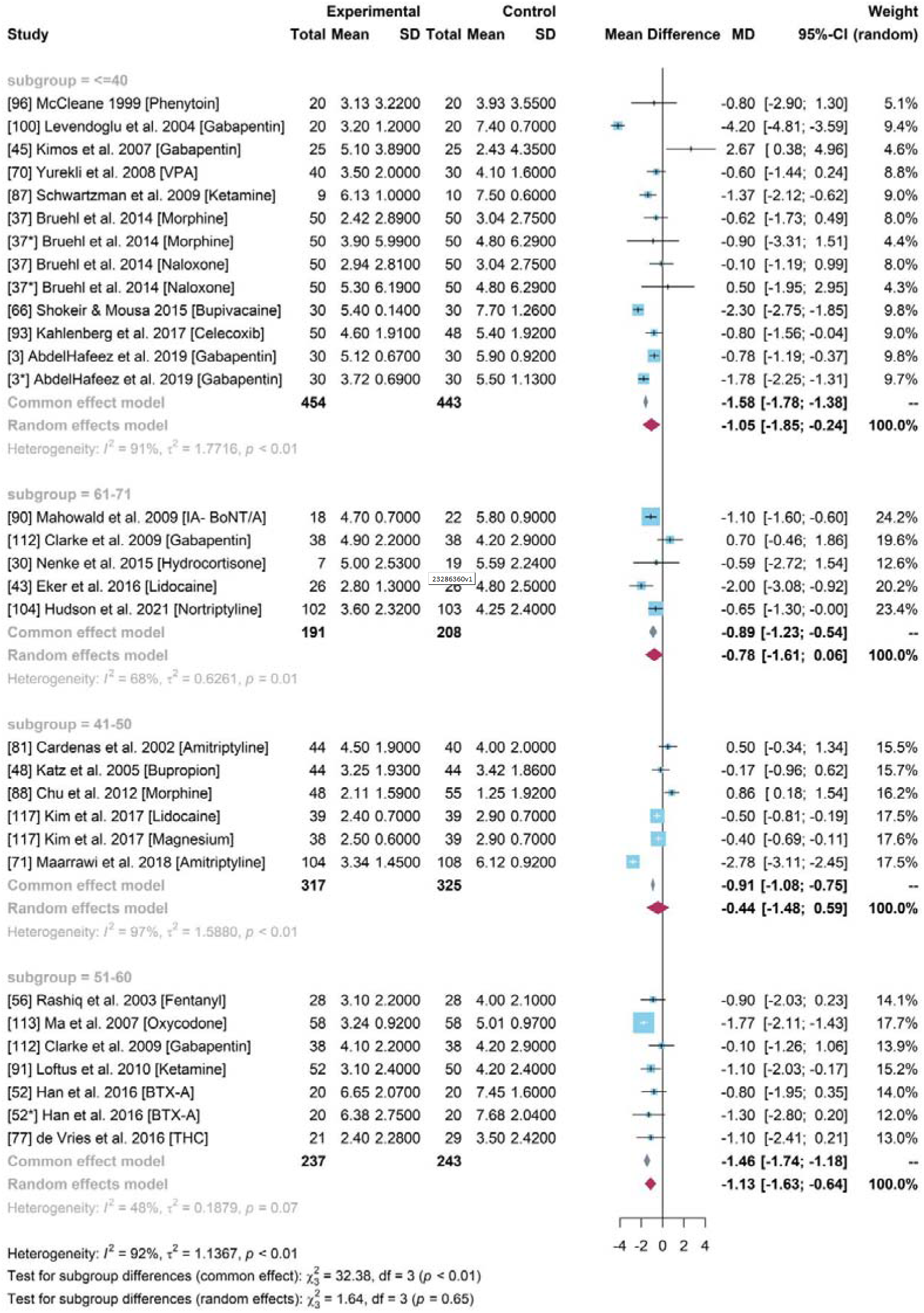
Forest plot for the mean difference of pain scores between experimental group and control group across different mean age of participants.

#### Subgroup analysis for pain ccore difference based on different age groups

It showed that the heterogeneity among studies with participants who were older than 50 years old had changed with decreased *I* ^2^ (*I* ^2^ = 48% for “51-60”, *I* ^2^ = 68% for “61-71”). A common effects model was chosen for subgroup “51-60”, which produced a higher estimation of pain reduction with a mean difference of -1.46 (95% CI = [-1.74, -1.18]). Based on the high heterogeneity (*I*^2^ > 50%), random effects models were built for other subgroups. The group with participants younger than 40 years older obtained a significant drug efficiency (MD = -1.05, 95% CI = [-1.85, -0.24]). The pooled drug effects [Figure 8] in the 41-50 and 61-71 years of age groups were much lower than the overall treatment effect of NSAID drugs identified in the PMA. The 95% CI of 0 indicated statistically ineffective compared to the placebo. The random effects models showed the decrease of heterogeneity indicating that age may be a source of heterogeneity.

### Sensitivity Analysis

The sensitivity analysis was conducted [Figure 12] for the PMA where some studies influenced the pooled results compared to the overall estimation (−0.89). To test this theory, study number 71 and 100 were omitted and the pooled results were much lower, -0.82 and - 0.79, respectively. Studies with *Amitriptyline* and *Gabapentin* produced unstable treatment results, and the absence of these showed an overestimation (study 81, 45) or underestimates (study 71, 100). Collectively, the high heterogeneity (*I*^2^= 92%, p-value < 0.01) was stable and a robust treatment effect with negative mean differences and a significant 95% CI remained. Therefore, the pooled treatment effects identified was credible.

**Figure 12.**
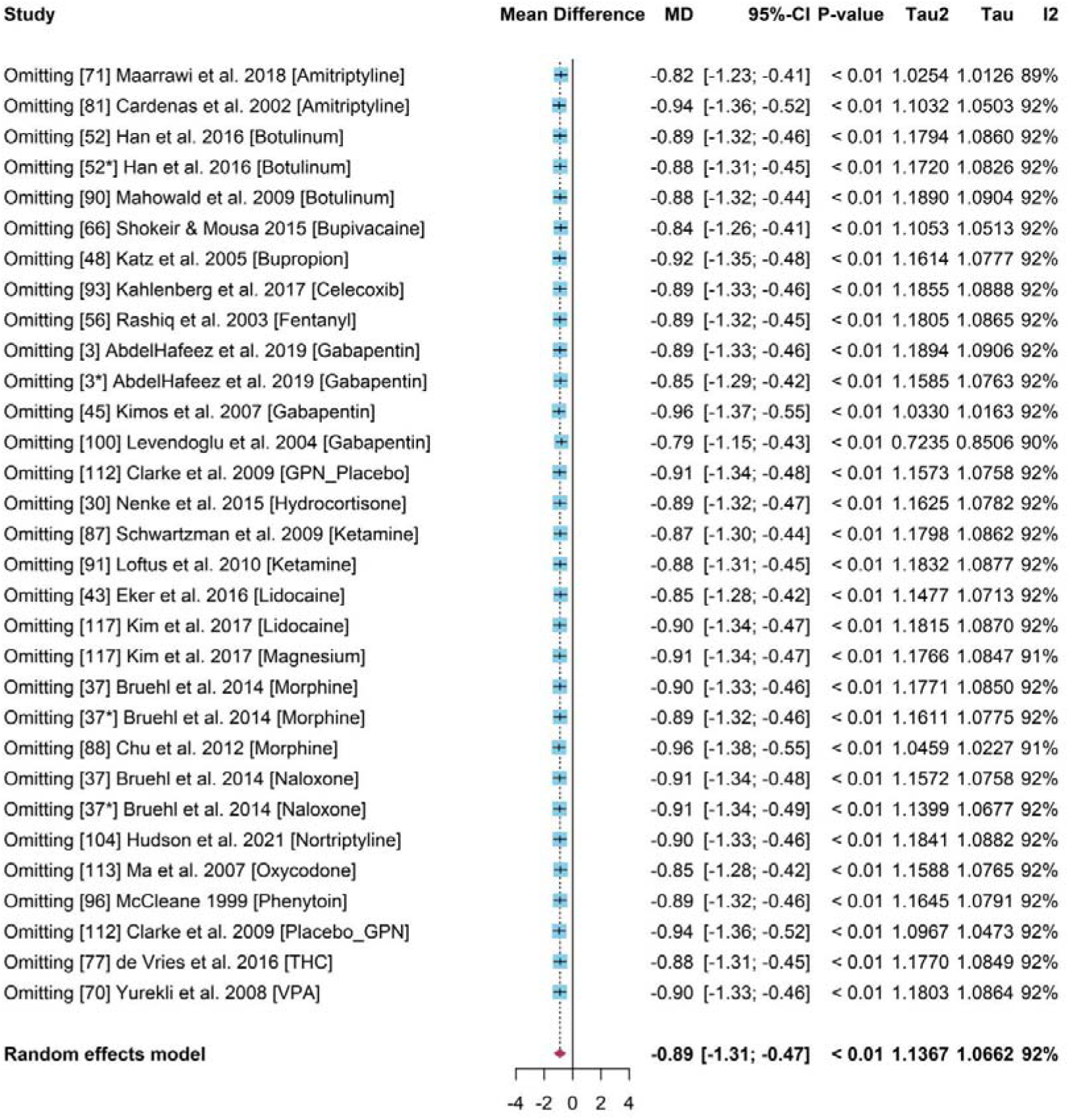
Forest plot for Sensitivity Analysis with studies in MA

### Publication bias

The funnel plots [Figure 13] within the PMA indicated symmetry. Although several studies were not within the remit of the funnel, the Egger’s test showed a p value (0.22) larger than 0.05 which indicated the lack of small-study effects.

**Figure 13.**
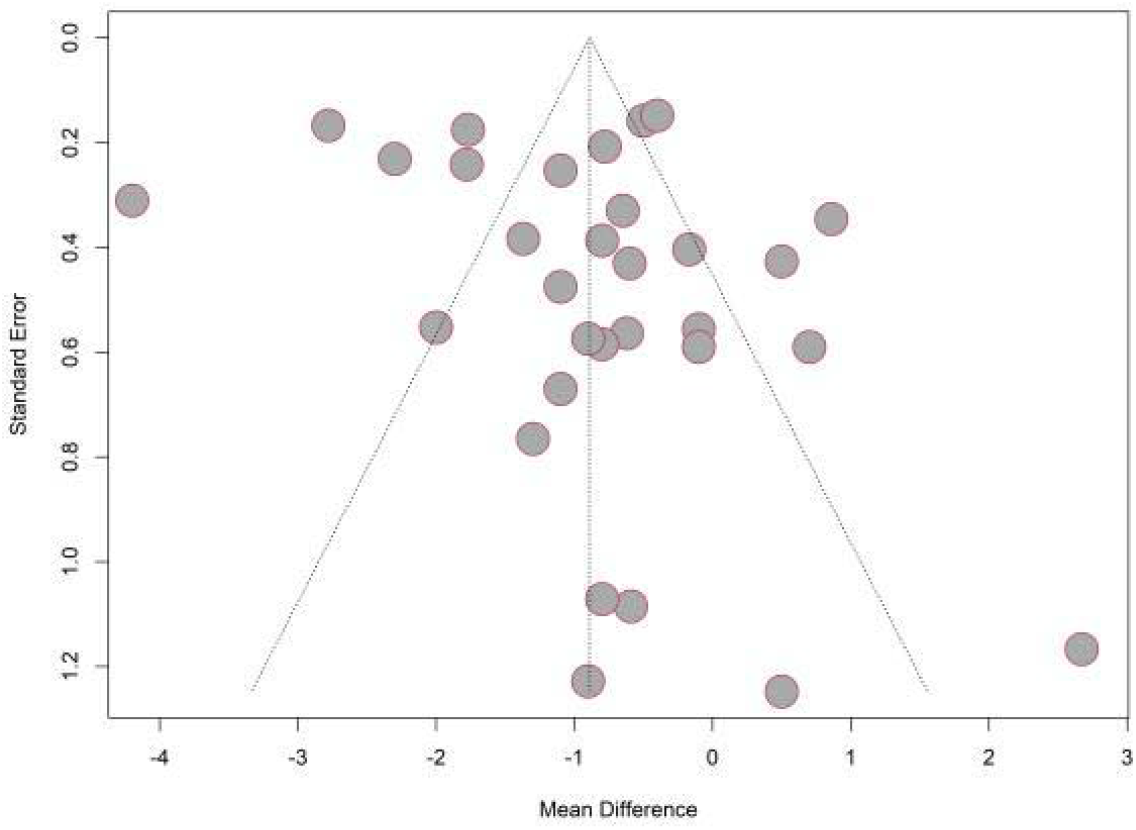
Funnel plot for studies used in PMA

## Discussion

We identified opiods and non-opiods were the two primary classes of pharmacological interventions in chronic pain management. The long-term use of opioids in the management of chronic non-malignant pain has come under scrutiny over the last few years and is now recommended that it should be offered only if benefits of initiating treatment would significantly outweigh the potential risks, and possibly as an adjunct to the primary intervention^[30]^. Our study has shown that judicious use of non-opioid medications along with other treatment modalities could provide better outcomes in managing chronic pain thereby removing long-term side-effects observed during opioid therapy. Opioids are widely used in the management of cancer pain and also non-cancer associated pain ^[25,26]^ However, recent evidence also indicates that opioids provide limited advantage in managing chronic non-cancer pain and have similar efficacy to non-opioids.^[29]^ The awareness of an opioid crisis globally has prompted clinicians to exercise caution in their prescription habits, but the WHO supports the use of opioids including Fentanyl and Methadone as an essential class of medication for the management of cancer pain^[27,28]^.

The meta-analysis of baseline pain scores lacked statistical significance between experimental and control groups. The significant reduction in chronic pain scores of patients taking NSAID versus non-steroidal opioid drugs compared to patients given placebo under a random effects model. The presence of a significant drug efficiency with *BTX-A* and *Ketamine* in interesting although the pooled results of other drugs and interventions had statistically insignificant results with a 95% CI of 0. The pooled evidence indicated Ketamine showed the highest pain reduction (1.26) followed by BTX-A (0.98). Studies testing on other drugs including Amitriptyline, Gabapentin, Morphine and Lidocaine had a high heterogeneity and insignificant drug efficiency. Overall, evidence from the PMA showed a strong efficacy within the NSAIDs group with managing pain which were remarkably narrowed when exclusive trials with low risk of bias were included^[11]^.

Many cancer patients are increasingly being cured or achieve long term remission, and prolonged use of opioids could result in aberrant behaviour and dependence, thus limiting opioid use to patients with a terminal illness or as part of end-of-life care. The CDC recommends the use of non-pharmacological therapies and non-opioid pharmacotherapy as the primary intervention in patients with chronic pain, and opioids should be offered only if benefits of initiating treatment significantly outweigh the potential risks and if possible, these should be used as an adjunct to the primary intervention.^[30].^ In this study, a pairwise meta-analysis and NMA consolidating the evidence of 46 studies was carried out, with the former comparing several different opioids. Morphine has traditionally been used for the management of moderate to severe chronic pain.^[33]^ Despite morphine being a potent analgesic [MD = 0.01 (95% CI=[-1.18,1.21], newer opioids are now being employed owing to their superior safety profile. Oxycodone and Fentanyl appear to be popular due to better availability and vast clinical experience including the well accepted effectiveness demonstrated, as per patient and clinically reported outcomes. Our results are aligned to these trends where the effectiveness is shown to include a MD 1.77 (95% CI=[-2.11,-1.43]) for Oxycodone and a MD of -0.90 (95% CI=[-2.03,0.23])] for Fentanyl.^[32]^ However, untoward gastrointestinal effects (constipation, nausea, and vomiting) still remain a major concern with opioid use and are often responsible for discontinuation of treatment.^[34,35]^ Recent evidence favours the use of a combination of oxycodone and naloxone in patients with chronic pain (after ensuring that there is no cause for porto-systemic anastomosis), to offer an improved bowel function without any effective change in analgesia.^[36].^ The concerns of developing tolerance, opioid-induced hyperalgesia, aberrant behaviour and dependence with opioids is a pragmatic reason to develop effective alternative treatment modalities especially for vulnerable individuals. In pairwise comparison, we observed Ketamine to be superior to other pharmacological interventions with a mean difference MD -1.26 (95% CI=[-1.85,-0.68]).

There are several guidelines recommending the use of Pregabalin, Gabapentin, Duloxetine, and Amitriptyline as first line drugs in the management of neuropathic pain ^[37]^. However, the use of gabapentinoids is being challenged as it lacks favourable robust evidence for efficacy against pain syndromes other than fibromyalgia, post herpetic neuralgia and diabetic neuropathy, and many clinicians have also highlighted the potential for misuse and developing dependence^[38,39,40]^. The use of BTX-A, Ketamine, Ningmitai and THC for the management of various chronic pain conditions is popular and well established^[12,13,14,15]^ and our study shows the effective use of these as analgesics when compared to placebo. There is evidence to support the efficacy of BTX-A for the management of neuropathic pain although the sample sizes used in the studies were small and therefore the real-world applicability remains limited^[16]^. BTX-A is also used in management of myofascial pains^[17,18]^ although further evidence on the efficacy and tolerability within all populations, especially those with existing co-morbidities needs to be evaluated. Ketamine was found to be beneficial in managing some neuropathic pains^[19]^ and as an infusion the rates of serious adverse effects were found to be similar to placebo.^[20] 3,24]^ Further studies are required to gather evidence to better understand its psychedelic effects and its role in the management of PTSD, anxiety and depression. A renewed use of magnesium in managing chronic pain has been demonstrated in some literature.^[50]^ Our results indicate similar evidence in the use of magnesium, but will require further research to determine the efficacy, safety and effectiveness in managing short, medium and long-term pain.

The NMA provided more reliable results with direct and indirect comparisons between different drugs under different study designs. However, only a small number of multi-arm trials were eligible and the distribution of trials studying different drugs was uneven. It resulted in the lack of direct evidence of certain drugs and their relative efficacy in the network was unstable due to excessive reliance on indirect comparisons. Therefore, well designed and robust clinical trials should be conducted to verify the efficacy of pharmaceutical interventions used in chronic pain management.

## Data Availability

All data produced in the present work are contained in the manuscript

## Conclusion

To the best of our knowledge, this is the first pairwise MA and NMA reporting the synthesis of the prevalence of the efficacy of pharmacological treatments used in the management of chronic pain with a large sample size of 17,708 participants. Management of long-term chronic pain needs to be prioritised for several reasons including humanitarian, the strain on the healthcare systems and the impact on the economy due to loss of productivity. The use of pharmaceutical agents in the long-term management of chronic pain has been debated for several decades, yet there has not been a consensus on this matter. This study supports the importance of generating better evidence by way of robust clinical trials, the need for drafting clinical guidelines that is pragmatic, practical as well as clinically significant and the use of better data-connectivity methods to improve clinical practice in the real-world.

## Appendix

**Table 6.**
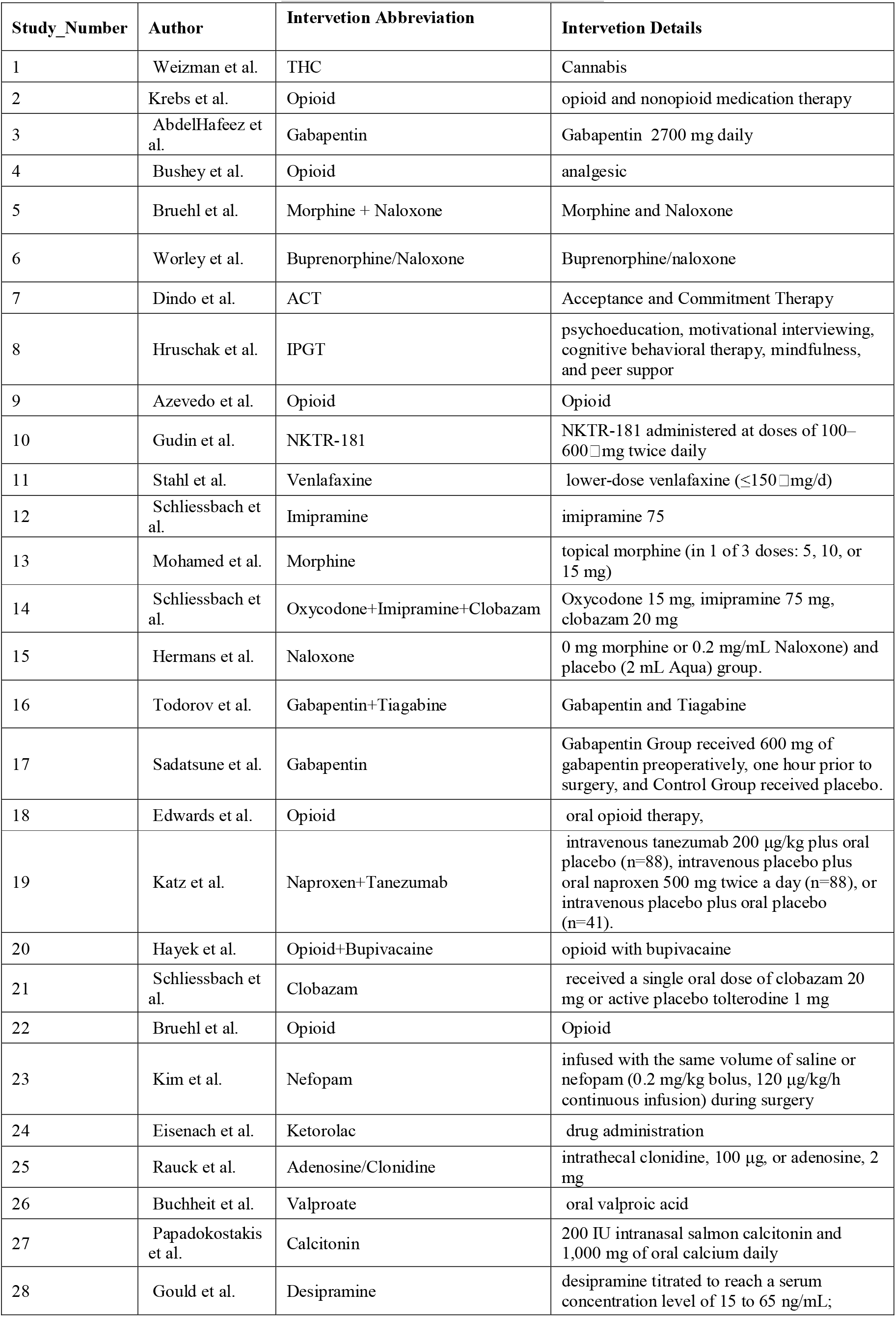
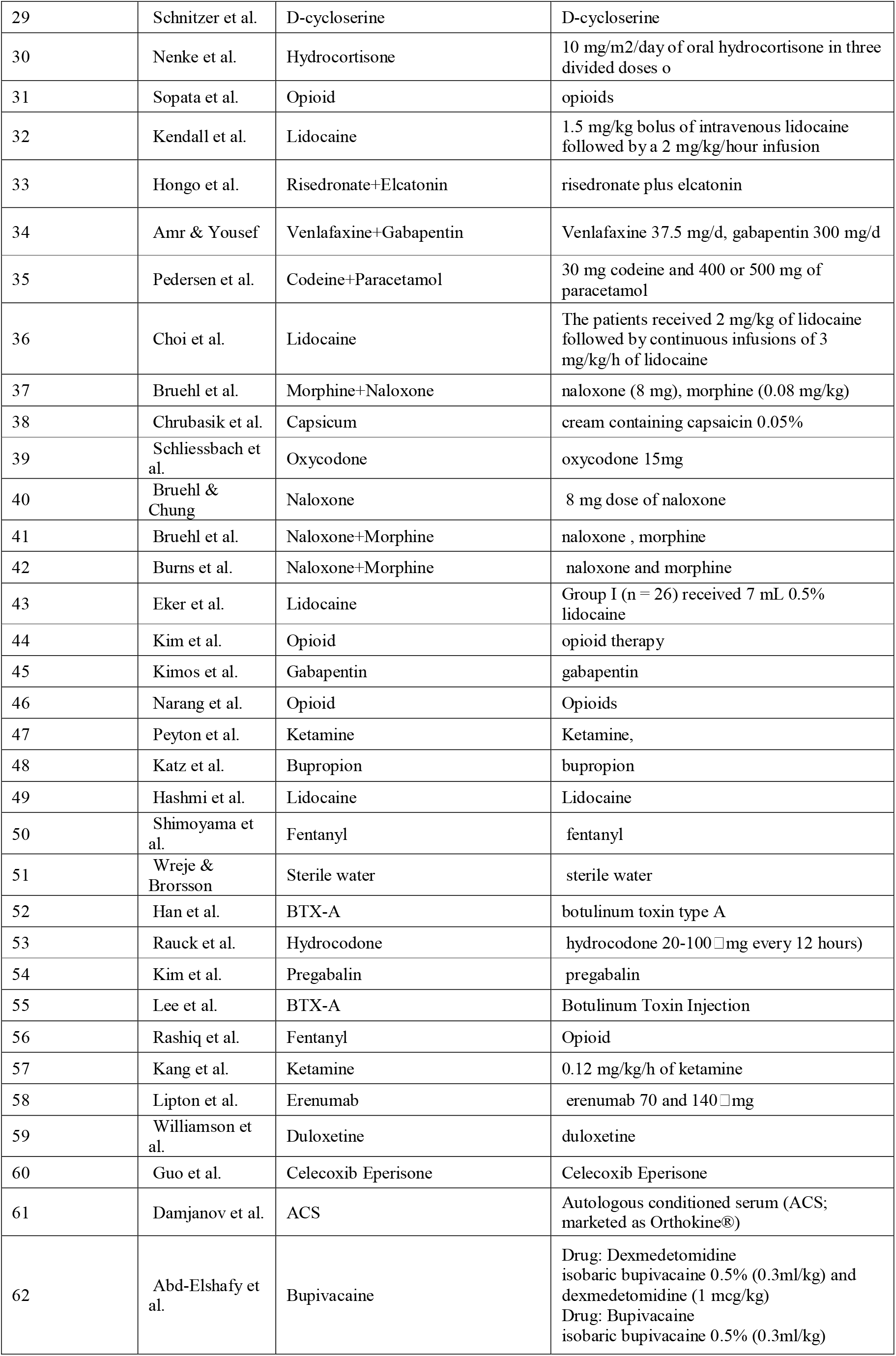
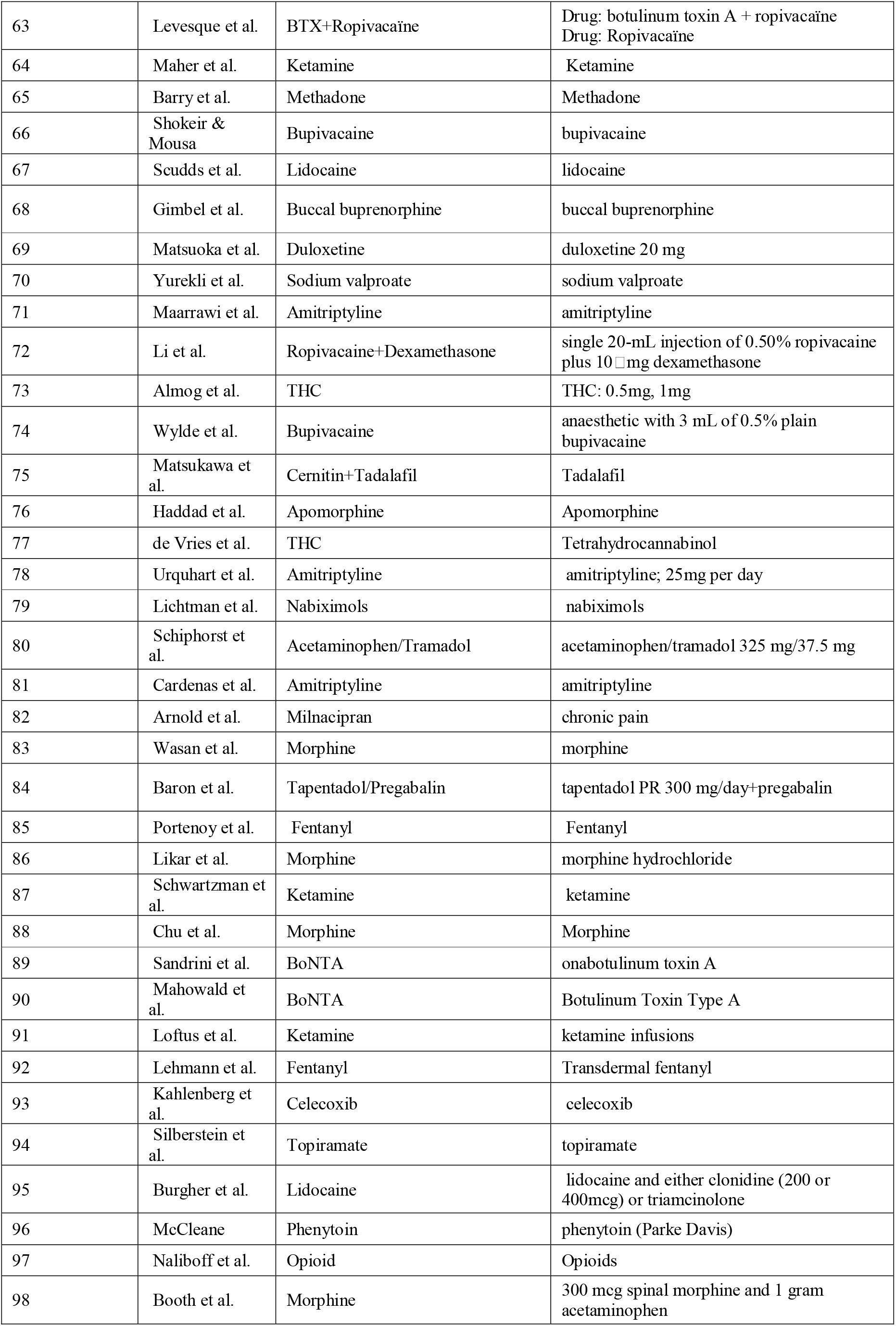
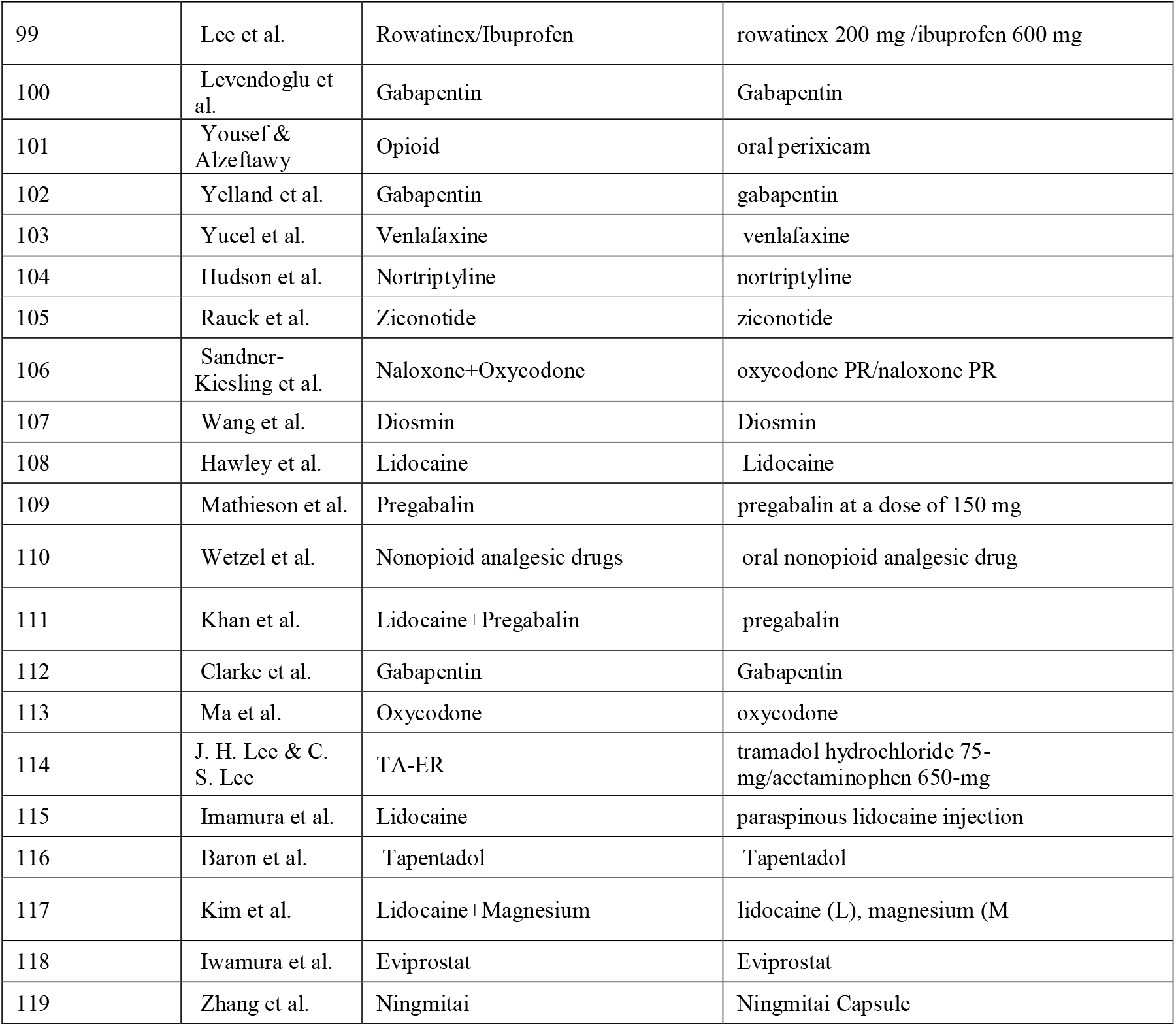
Intervetions used in studies

## Notes

**Conflicts of interest** AS has received funding from Neurokor, Boston Scientific and Medtronic. PP has received research grants from Novo Nordisk, Queen Mary University of London, John Wiley & Sons, Otsuka, outside the submitted work. All other authors report no conflict of interest. The views expressed are those of the authors and not necessarily those of the NHS, the National Institute for Health Research, the Department of Health and Social Care or the Academic institutions.

### Competing Interest Statement

The authors have declared no competing interest.

### Funding Statement

This study did not receive any funding

### Author Declarations

The source data is available in the public domain and can be viewed using search engines such as Pubmed, Science direct etc.

## References

1. Dahlhamer J, Lucas J, Zelaya, C, et al. Prevalence of Chronic Pain and High-Impact Chronic Pain Among Adults — United States, 2016. MMWR Morb Mortal Wkly Rep 2018;67:1001–1006. DOI: http://dx.doi.org/10.15585/mmwr.mm6736a2externalicon

2. Salanti G. Indirect and mixed-treatment comparison, network, or multiple-treatments meta-analysis: many names, many benefits, many concerns for the next generation evidence synthesis tool. Res Synth Methods. 2012 Jun;3(2):80–97. doi: 10.1002/jrsm.1037. Epub 2012 Jun 11. PMID: 26062083.

3. Brown, C.A., Lilford, R.J The stepped wedge trial design: a systematic review. BMC Med Res Methodol 6, 54 (2006). https://doi.org/10.1186/1471-2288-6-54

4. Lumley, T. “Network Meta-analysis for Indirect Treatment Comparisons.” Statistics in Medicine 21 (2002): null. https://doi.org/10.1002/SIM.1201.

5. Li, T., Puhan, M.A., Vedula, S.S et al. Network meta-analysis-highly attractive but more methodological research is needed. BMC Med 9, 79 (2011). https://doi.org/10.1186/1741-7015-9-79

6. Caldwell, D., A. Ades, and J. Higgins. “Simultaneous Comparison of Multiple Treatments: Combining Direct and Indirect Evidence.” BMJ: British Medical Journal 331 (2005): 897–900. https://doi.org/10.1136/bmj.331.7521.897.

7. Jansen JP, Fleurence R, Devine B, Itzler R, Barrett A, Hawkins N, Lee K, Boersma C, Annemans L, Cappelleri JC. Interpreting indirect treatment comparisons and network meta-analysis for health-care decision making: report of the ISPOR task force on indirect treatment comparisons good research practices: part 1. Value Health. 2011;14(4):417–28.

8. Efthimiou O, Debray TP, van Valkenhoef G, Trelle S, Panayidou K, Moons KG, Reitsma JB, Shang A, Salanti G. GetReal in network meta-analysis: a review of the methodology. Res Synth Methods. 2016;7(3):236–63.

9. Dallenbach KM. Pain: History and present status. Am J Psychol 1939; 52: 331.

10. Turk DC and Monarch ES. Biopsychosocial perspective on chronic pain. In: Turk DC, Gatchel RJ, eds. Psychological Approaches to Pain Management: A Practitioner’s Handbook. 2nd ed. Guilford. New York. 2002..

11. Brown, C.A., Lilford, R.J The stepped wedge trial design: a systematic review. BMC Med Res Methodol 6, 54 (2006). https://doi.org/10.1186/1471-2288-6-54

12. Campbell, J. (1996, November 11). Presidential Address. Speech given at the American Pain Society, Washington, DC..

13. Zimmer Z, Fraser K, Grol-Prokopczyk H, Zajacova A. A global study of pain prevalence across 52 countries: examining the role of country-level contextual factors: Examining the role of country-level contextual factors. Pain 2022; 163: 1740–50.

14. Relieving pain in America: A blueprint for transforming prevention, care, education, and research. Washington, D.C.: National Academies Press, 2011.

15. Caldwell, D., A. Ades, and J. Higgins. “Simultaneous Comparison of Multiple Treatments: Combining Direct and Indirect Evidence.” BMJ: British Medical Journal 331 (2005): 897–900. https://doi.org/10.1136/bmj.331.7521.897.

16. U.S. Department of Health and Human Services (2019, May). Pain Management Best Practices Inter-Agency Task Force Report: Updates, Gaps, Inconsistencies, and Recommendations. Retrieved from U. S. Department of Health and Human Services website: https://www.hhs.gov/ash/advisory-committees/pain/reports/index.html..

17. Borenstein M, Hedges LV, Higgins JP, Rothstein HR (2010): A basic introduction to fixed-effect and random-effects models for meta-analysis. Research Synthesis Methods, 1, 97–111

18. Rothstein, H.R., Sutton, A.J., Borenstein, M.: Publication Bias in Meta Analysis: Prevention, Assessment and Adjustments. Wiley, Chichester (2005)

19. Béliveau A, Boyne DJ, Slater J, Brenner D, Arora P. BUGSnet: an R package to facilitate the conduct and reporting of Bayesian network Meta-analyses. BMC Med Res Methodol. 2019 Oct 22;19(1):196. doi: 10.1186/s12874-019-0829-2. PMID: 31640567; PMCID: PMC6805536.

20. Jansen JP, Fleurence R, Devine B, Itzler R, Barrett A, Hawkins N, Lee K, Boersma C, Annemans L, Cappelleri JC. Interpreting indirect treatment comparisons and network meta-analysis for health-care decision making: report of the ISPOR task force on indirect treatment comparisons good research practices: part 1. Value Health. 2011;14(4):417–28.

21. Opioid overdose. Who.intlJ: www.who.int/news-room/fact-sheets/detail/opioid-overdose..

22. Ho KY, Cardosa MS, Chaiamnuay S, et al. Practice advisory on the appropriate use of NSAIDs in primary care. J Pain Res 2020; 13: 1925–39.

23. Chung JW, Zeng Y, Wong TK. Drug therapy for the treatment of chronic nonspecific low back pain: systematic review and meta-analysis. Pain Physician 2013; 16: E685–704.

24. Laine L, Curtis SP, Cryer B, Kaur A, Cannon CP, MEDAL Steering Committee. Assessment of upper gastrointestinal safety of etoricoxib and diclofenac in patients with osteoarthritis and rheumatoid arthritis in the Multinational Etoricoxib and Diclofenac Arthritis Long-term (MEDAL) programme: a randomised comparison. Lancet 2007; 369: 465–73.

25. Griffin MR. High-dose non-steroidal anti-inflammatories: painful choices. Lancet 2013; 382: 746–8.

26. Trelle S, Reichenbach S, Wandel S, et al. Cardiovascular safety of non-steroidal anti-inflammatory drugs: network meta-analysis. BMJ 2011; 342: c7086.

27. da Costa BR, Pereira TV, Saadat P, et al. Effectiveness and safety of non-steroidal anti-inflammatory drugs and opioid treatment for knee and hip osteoarthritis: network meta-analysis. BMJ 2021; 375: 2321.

28. Overview | Chronic pain (primary and secondary) in over 16s: assessment of all chronic pain and management of chronic primary pain | Guidance | NICE. https://www.nice.org.uk/guidance/ng193 x(accessed Dec 9, 2022).

29. Eccleston C, Aldington D, Moore A, de C Williams AC. Pragmatic but flawed: the NICE guideline on chronic pain. Lancet 2021; 397: 2029–31.

30. Wood H. Anti-inflammatory drugs could cause chronic pain. Nat Rev Neurol 2022; 18: 382.

31. Bannuru RR, Osani MC, Vaysbrot EE, et al. OARSI guidelines for the non-surgical management of knee, hip, and polyarticular osteoarthritis. Osteoarthritis Cartilage 2019; 27: 1578–89.

32. Enthoven WTM, Roelofs PDDM, Deyo RA, van Tulder MW, Koes BW. Non-steroidal anti-inflammatory drugs for chronic low back pain. Cochrane Database Syst Rev 2016; 2: CD012087.

33. Hary V, Schitter S, Martinez V. Efficacy and safety of botulinum A toxin for the treatment of chronic peripheral neuropathic pain: A systematic review of randomized controlled trials and meta-analysis. Eur J Pain 2022; 26: 980–90.

34. Orhurhu V, Orhurhu MS, Bhatia A, Cohen SP. Ketamine infusions for chronic pain: A systematic review and meta-analysis of randomized controlled trials. Anesth Analg 2019; 129: 241–54.

35. Jin C, Chen Z, Zhang J. Meta-analysis of the efficacy of Ningmitai capsule on the treatment of chronic prostatitis in China. Medicine (Baltimore) 2018; 97: e11840.

36. Whiting PF, Wolff RF, Deshpande S, et al. Cannabinoids for medical use: A systematic review and meta-analysis: A systematic review and meta-analysis. JAMA 2015; 313: 2456–73.

37. Wei J, Zhu X, Yang G, et al. The efficacy and safety of botulinum toxin type A in treatment of trigeminal neuralgia and peripheral neuropathic pain: A meta-analysis of randomized controlled trials. Brain Behav 2019; 9: e01409.

38. Khalifeh M, Mehta K, Varguise N, Suarez-Durall P, Enciso R. Botulinum toxin type A for the treatment of head and neck chronic myofascial pain syndrome. J Am Dent Assoc 2016; 147: 959-973.e1.

39. Meister MR, Brubaker A, Sutcliffe S, Lowder JL. Effectiveness of botulinum toxin for treatment of symptomatic pelvic floor myofascial pain in women: A systematic review and meta-analysis: A systematic review and meta-analysis. Female Pelvic Med Reconstr Surg 2021; 27: e152–60.

40. Guimarães Pereira JE, Ferreira Gomes Pereira L, Mercante Linhares R, Darcy Alves Bersot C, Aslanidis T, Ashmawi HA. Efficacy and safety of ketamine in the treatment of neuropathic pain: A systematic review and meta-analysis of randomized controlled trials. J Pain Res 2022; 15: 1011–37.

41. Cohen SP, Bhatia A, Buvanendran A, et al. Consensus guidelines on the use of intravenous ketamine infusions for chronic pain from the American society of regional anesthesia and pain medicine, the American academy of pain medicine, and the American society of anesthesiologists. Reg Anesth Pain Med 2018; : 1.

42. Zhang K, Liu Y, Yang W, et al. Efficacy and safety of Ningmitai capsule in patients with chronic prostatitis/chronic pelvic pain syndrome: A multicenter, randomized, double-blind, placebo-controlled trial. Urology 2021; 153: 264–9.

43. Jing Z, Liying G, Zhenqing W, et al. Efficacy and safety of Ningmitai capsules in patients with chronic epididymitis: A prospective, parallel randomized controlled clinical trial. Evid Based Complement Alternat Med 2021; 2021: 9752592.

44. Campbell G, Hall WD, Peacock A, et al. Effect of cannabis use in people with chronic non-cancer pain prescribed opioids: findings from a 4-year prospective cohort study. Lancet Public Health 2018; 3: e341–50.

45. Boland EG, Bennett MI, Allgar V, Boland JW. Cannabinoids for adult cancer-related pain: systematic review and meta-analysis. BMJ Support Palliat Care 2020; 10: 14–24.

46. Pergolizzi JV Jr, Magnusson P, Christo PJ, et al. Opioid therapy in cancer patients and survivors at risk of addiction, misuse or complex dependency. Front Pain Res (Lausanne) 2021; 2: 691720.

47. Boudreau D, Von Korff M, Rutter CM, et al. Trends in long-term opioid therapy for chronic non-cancer pain. Pharmacoepidemiol Drug Saf 2009; 18: 1166–75.

48. The Lancet Public Health. Opioid overdose crisis: time for a radical rethink. Lancet Public Health 2022; 7: e195.

49. World Health Organization Model List of Essential Medicines – 22nd List, 2021. Geneva: World Health Organization; 2021 (WHO/MHP/HPS/EML/2021.02)

50. Busse JW, Wang L, Kamaleldin M, et al. Opioids for chronic noncancer pain: A systematic review and meta-analysis: A systematic review and meta-analysis. JAMA 2018; 320: 2448–60.

51. Dowell D, Ragan KR, Jones CM, Baldwin GT, Chou R. CDC clinical practice guideline for Prescribing Opioids for pain - United States, 2022. MMWR Recomm Rep 2022; 71: 1–95.

52. Meng Z, Yu J, Acuff M, et al. Tolerability of opioid analgesia for chronic pain: A network meta-analysis. Sci Rep 2017; 7: 1995.

53. Caraceni A, Hanks G, Kaasa S, et al. Use of opioid analgesics in the treatment of cancer pain: evidence-based recommendations from the EAPC. Lancet Oncol 2012; 13: e58–68.

54. World Health Organization. Cancer pain relief, second edition, with a guide to opioid availability. Geneva: World Health Organization; 1996..

55. Noori A, Sadeghirad B, Wang L, et al. Comparative benefits and harms of individual opioids for chronic non-cancer pain: a systematic review and network meta-analysis of randomised trials. Br J Anaesth 2022; 129: 394–406.

56. Gehling M, Hermann B, Tryba M. Meta-analysis of dropout rates in randomized controlled clinical trials: opioid analgesia for osteoarthritis pain: Opioid analgesia for osteoarthritis pain. Schmerz 2011; 25: 296–305.

57. Huang L, Zhou J-G, Zhang Y, et al. Opioid-induced constipation relief from fixed-ratio combination prolonged-release oxycodone/naloxone compared with oxycodone and morphine for chronic nonmalignant pain: A systematic review and meta-analysis of randomized controlled trials. J Pain Symptom Manage 2017; 54: 737-748.e3.

58. Schlereth T. Guideline “diagnosis and non interventional therapy of neuropathic pain” of the German Society of Neurology (deutsche Gesellschaft für Neurologie). Neurol Res Pract 2020; 2: 16.

59. Wiffen PJ, Derry S, Bell RF, et al. Gabapentin for chronic neuropathic pain in adults. Cochrane Database Syst Rev 2017; 6: CD007938.

60. Shanthanna H, Gilron I, Rajarathinam M, et al. Benefits and safety of gabapentinoids in chronic low back pain: A systematic review and meta-analysis of randomized controlled trials. PLoS Med 2017; 14: e1002369.

61. Evoy KE, Morrison MD, Saklad SR. Abuse and misuse of pregabalin and gabapentin. Drugs 2017; 77: 403–26.

62. Alberti FF, Becker MW, Blatt CR, Ziegelmann PK, da Silva Dal Pizzol T, Pilger D. Comparative efficacy of amitriptyline, duloxetine and pregabalin for treating fibromyalgia in adults: an overview with network meta-analysis. Clin Rheumatol 2022; 41: 1965–78.

63. Urquhart DM, Wluka AE, van Tulder M, et al. Efficacy of low-dose amitriptyline for chronic low back pain: A randomized clinical trial: A randomized clinical trial. JAMA Intern Med 2018; 178: 1474–81.

64. Sankar V, Oommen AE, Thomas A, Nair JV, James JS. Efficacy, safety and cost effectiveness of amitriptyline and pregabalin in patients with diabetic peripheral neuropathy. Indian J Pharm Sci 2017; 79. DOI:10.4172/pharmaceutical-sciences.1000274.

65. Levene JL, Weinstein EJ, Cohen MS, et al. Local anesthetics and regional anesthesia versus conventional analgesia for preventing persistent postoperative pain in adults and children: A Cochrane systematic review and meta-analysis update. J Clin Anesth 2019; 55: 116–27.

66. Donado C, Lobo K, Velarde-Álvarez MF, et al. Continuous regional anesthesia and inpatient rehabilitation for pediatric complex regional pain syndrome. Reg Anesth Pain Med 2017; 42: 527–34.

67. Meng H, Fei Q, Wang B, et al. Epidural injections with or without steroids in managing chronic low back pain secondary to lumbar spinal stenosis: a meta-analysis of 13 randomized controlled trials. Drug Des Devel Ther 2015; 9: 4657–67.

68. Egli S, Pfister M, Ludin SM, Puente de la Vega K, Busato A, Fischer L. Long-term results of therapeutic local anesthesia (neural therapy) in 280 referred refractory chronic pain patients. BMC Complement Altern Med 2015; 15: 200.

69. Challapalli V, Tremont-Lukats IW, McNicol ED, Lau J, Carr DB. Systemic administration of local anesthetic agents to relieve neuropathic pain. Cochrane Database Syst Rev 2005; : CD003345.

70. Abd-Elshafy SK, Abdallal F, Kamel EZ, et al. Paravertebral dexmedetomidine in video-assisted thoracic surgeries for acute and chronic pain prevention. Pain Physician 2019; 22: 271–80.

71. Park R, Ho AM-H, Pickering G, Arendt-Nielsen L, Mohiuddin M, Gilron I. Efficacy and safety of magnesium for the management of chronic pain in adults: A systematic review: A systematic review. Anesth Analg 2020; 131: 764–75.

